# Ancestrally diverse genome-wide association analysis highlights ancestry-specific differences in genetic regulation of plasma protein levels

**DOI:** 10.1101/2024.09.27.24314500

**Authors:** Chloé Sarnowski, Jianzhong Ma, Ngoc Quynh H. Nguyen, Ron C Hoogeveen, Christie M Ballantyne, Josef Coresh, Alanna C Morrison, Nilanjan Chatterjee, Eric Boerwinkle, Bing Yu

## Abstract

Fully characterizing the genetic architecture of circulating proteins in multi-ancestry populations provides an unprecedented opportunity to gain insights into the etiology of complex diseases. We characterized and contrasted the genetic associations of plasma proteomes in 9,455 participants of European and African (19.8%) ancestry from the Atherosclerosis Risk in Communities Study. Of 4,651 proteins, 1,408 and 2,565 proteins had protein-quantitative trait loci (pQTLs) identified in African and European ancestry respectively, and twelve unreported potentially causal protein-disease relationships were identified. Shared pQTLs across the two ancestries were detected in 1,113 aptamer-region pairs pQTLs, where 53 of them were not previously reported (all *trans* pQTLs). Sixteen unique protein-cardiovascular trait pairs were colocalized in both European and African ancestry with the same candidate causal variants. Our systematic cross-ancestry comparison provided a reliable set of pQTLs, highlighted the shared and distinct genetic architecture of proteome in two ancestries, and demonstrated possible biological mechanisms underlying complex diseases.

## Introduction

Human proteins, collectively referred as the proteome, play key roles in numerous biological processes, such as mediating homeostasis via signaling, immune responses, nutrient transport, and tissue remodeling. Along with other circulating factors, proteins are believed to directly contribute to disease onset and progression and other complex processes such as aging and fetal development. (1) Characterization of the genetic regulation of proteins can help bridge the gap between the genome and complex human diseases and provide further etiological insights and novel therapeutic opportunities. (1,2)

Proteins lie in the central layer of information from the genome to the phenome. Recent studies have started to build a genomic atlas of the human proteome via genome-wide association studies (GWASs) on the scale of proteome for the protein levels measured using the aptamer-based SomaScan or the antibody-based Olink technologies. (1–4) Identification of protein-quantitative trait loci (pQTLs) can provide insights into disease mechanisms and shared genetic architecture across diseases. Integrating pQTLs and disease-variant associations identified through GWASs as done by recent proteome-wide MR studies (5–7) provide an opportunity to identify causal pathways and thus help elucidating how proteins mediate genetic risk to disease onset and point to potential drug targets. (3,4)

With few exceptions, studies aiming at integrating the human proteome and genome have focused on participants of European ancestry, (1–4,8,9) reminiscent of early GWAS for diseases and other traits. (10) The genetic architecture of proteome in East Asian, South Asian, or African populations is not well characterized. (11,12) As the patterns of genetic variation among populations have impacts on both disease risk and treatment efficacy and safety, it is crucial to extend the effort of mapping the proteome and genome in relation to human disease to more ancestrally diverse populations. A study leveraging proteomic profiles of ∼1,300 proteins in ∼1,850 adults of African ancestry from the Jackson Heart Study (JHS) identified novel genetic determinants of circulating proteins, many of which are important in cardiovascular diseases. (13) A recent study performed in 2,958 Han Chinese participants validated 62% of the pQTL associations in European populations. (12) Another recent study conducted in the UK Biobank (8) investigated ancestry-specific pQTLs within five population groups non-primarily of European descent and identified a higher allelic enrichment in African ancestry participants (n=931) driving associations that were not detected in European ancestry participants. In addition, this population group was the one with the lowest validation rate of primary pQTLs in the European ancestry pQTLs findings (68% *versus* 97% for the other population groups). It is thus important to understand to what extent the established genomic atlas of the proteome and the resultant insights into the disease mechanisms explained by the mediation of proteins are shared across ancestries.

Understanding the shared and distinct genetic effect on circulating proteins in African and European ancestries may increase the precision of using genomic information for clinical practice. (14) In addition, identification of the shared genetic variants associated with both the level of proteins and diseases allows for generating immediate hypothesis for experimental validation on the potential causal genes and/or genetic variants of diseases by providing instrument variables to study the causal relationship between the protein and disease. We report here a GWAS of proteomic data (4,877 plasma proteins measured using SomaScan) conducted in 7,584 European (EA) and 1,871 African (AA) ancestry participants from the Atherosclerosis Risk in Communities (ARIC) study, characterizing whether both the proximal (cis) and distal (trans) pQTLs identified are shared across the two ancestral groups, using a larger number of proteins and larger sample size compared to previous pQTL studies conducted in AA participants. We further leverage the pQTLs identified, along with expression QTLs and disease GWAS summary statistics, to identify shared genetic variants and establish potential causal relationships between circulating proteins and complex diseases and traits using colocalization and Mendelian Randomization techniques. A flowchart detailing our analysis strategy is presented in **Figure 1**.

**Figure 1.**
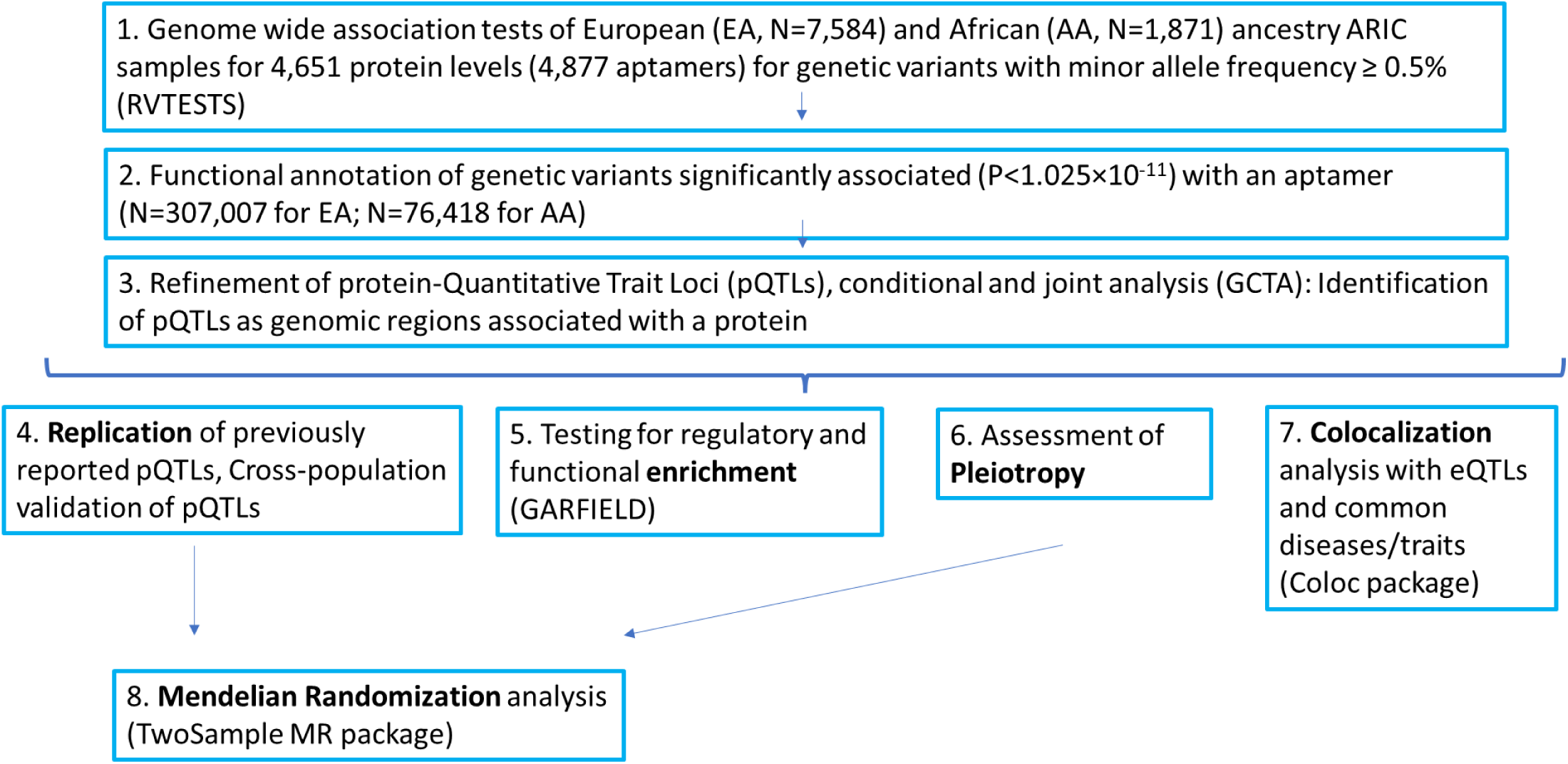
Flowchart displaying the strategy for the protein Quantitative Trait Loci (pQTLs) analysis in ARIC.

## Results

### Genetic determinants of plasma proteome in African and European ancestry participants

We performed a GWAS of 18.1 million variants with minor allele frequency (MAF) ≥ 0.5% among 1,871 AA participants (1,156 women, 61.8%) and 9.7 million variants with MAF ≥ 0.5% among 7,584 EA participants (3,964 women, 52.3%) from the ARIC study using an additive genetic model on 4,877 plasma aptamers (4,651 proteins). A description of the ARIC participants included in the genetic proteomic analyses is presented in **Supplementary Table 1**. We identified 121,996 and 804,783 associated pairs of variant and aptamer at P<1.025×10^-11^ in AA and EA, respectively. The genomic inflation factor ranged from 0.987 to 1.029 and 0.985 to 1.041 with a mean of 1.006 and 1.007 for AA and EA, respectively, indicating well-controlled type I error and population stratification.

The identified associations involved 1,441 and 2,648 aptamers in AA and EA, respectively, representing 1,408 (AA) and 2,565 (EA) proteins, and 76,418 (AA) and 307,007 (EA) unique variants. As many of the adjacent variants associated with an aptamer may simply represent a single signal due to linkage disequilibrium (LD), we grouped the genome-wide significant variants into one or more associated regions to identify conditionally independent and non-overlapping associations (**Methods**).

After conditional analysis, we identified 1,746 significant aptamer-region pairs associations (37% cis) among 807 regions and 1,408 proteins in AA (**Supplementary Tables 2** and **3**, **Figure 2**) and 4,315 significant associations (24% cis) among 1,490 regions and 2,565 proteins in EA (**Supplementary Tables 2** and **4**, **Figure 2**). The breakdown of cis versus trans associations for the sentinel variant to protein associations or genomic regions or proteins is provided in **Supplementary Table 2**. Of the 1,746 and 4,315 pQTLs identified in AA and EA, 284 (16%) and 834 (19%) respectively had multiple conditionally significant associations, of which 230/530 were in cis (**Figure 3**). Conditional analyses yielded 2,160 pairs of conditionally independent aptamer-variants associations in AA and 5,901 in EA (**Supplementary Tables 5** and **6**).

**Figure 2.**
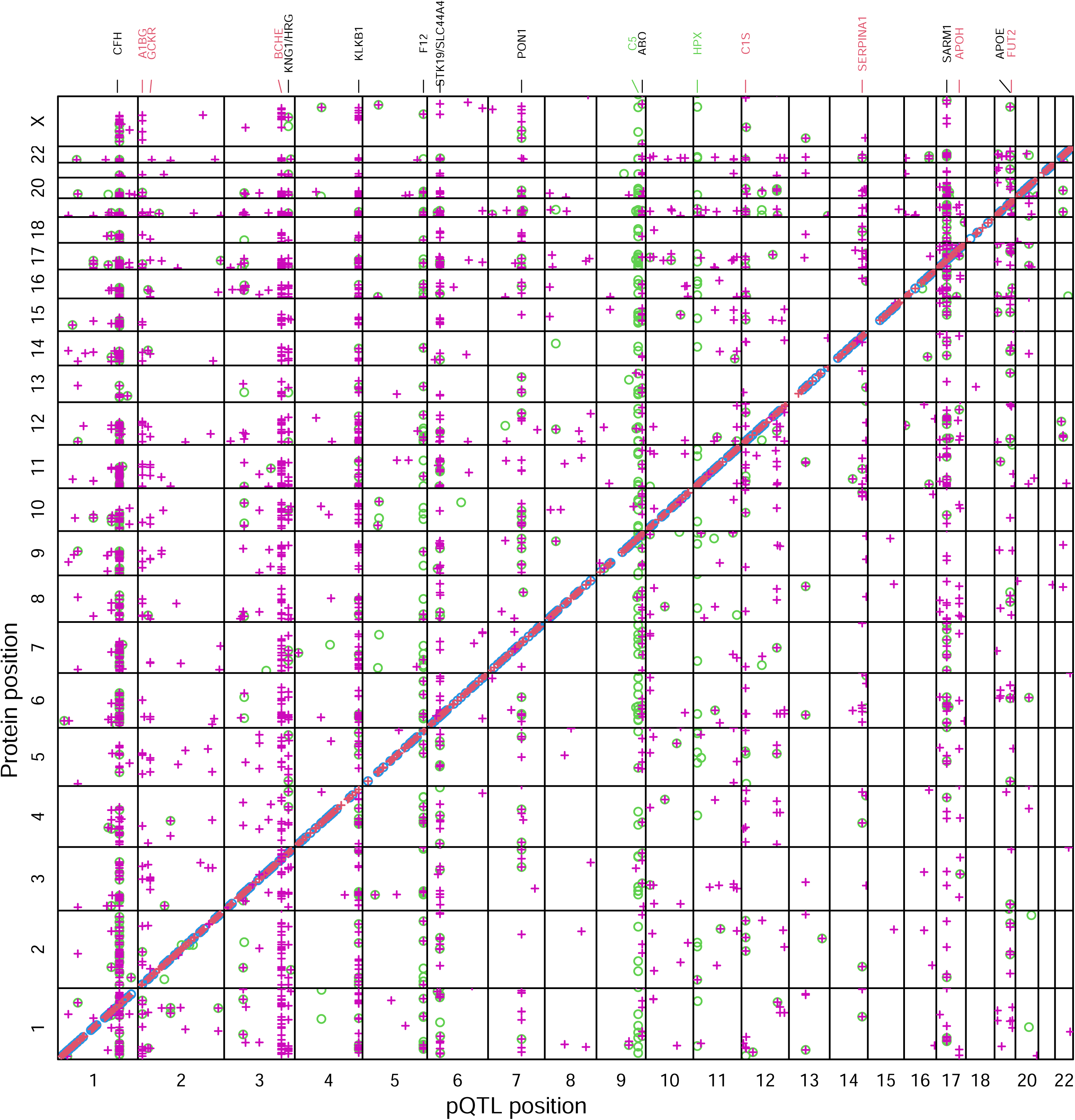
Genomic locations of protein Quantitative Trait Loci (pQTLs) European ancestry (EA) signals are represented by a “+” symbol with cis signals represented in red and trans signals in magenta. African ancestry (AA) signals are represented by a “o” symbol with cis signals represented in blue and trans signals in green. The x axis indicates the positions of the sentinel variants of the pQTLs, and the y axis indicates the genomic locations of the coding genes of the proteins. Highly pleiotropic genomic regions are annotated at the top (red: EA only; green: AA only; black: both EA and AA).

**Figure 3.**
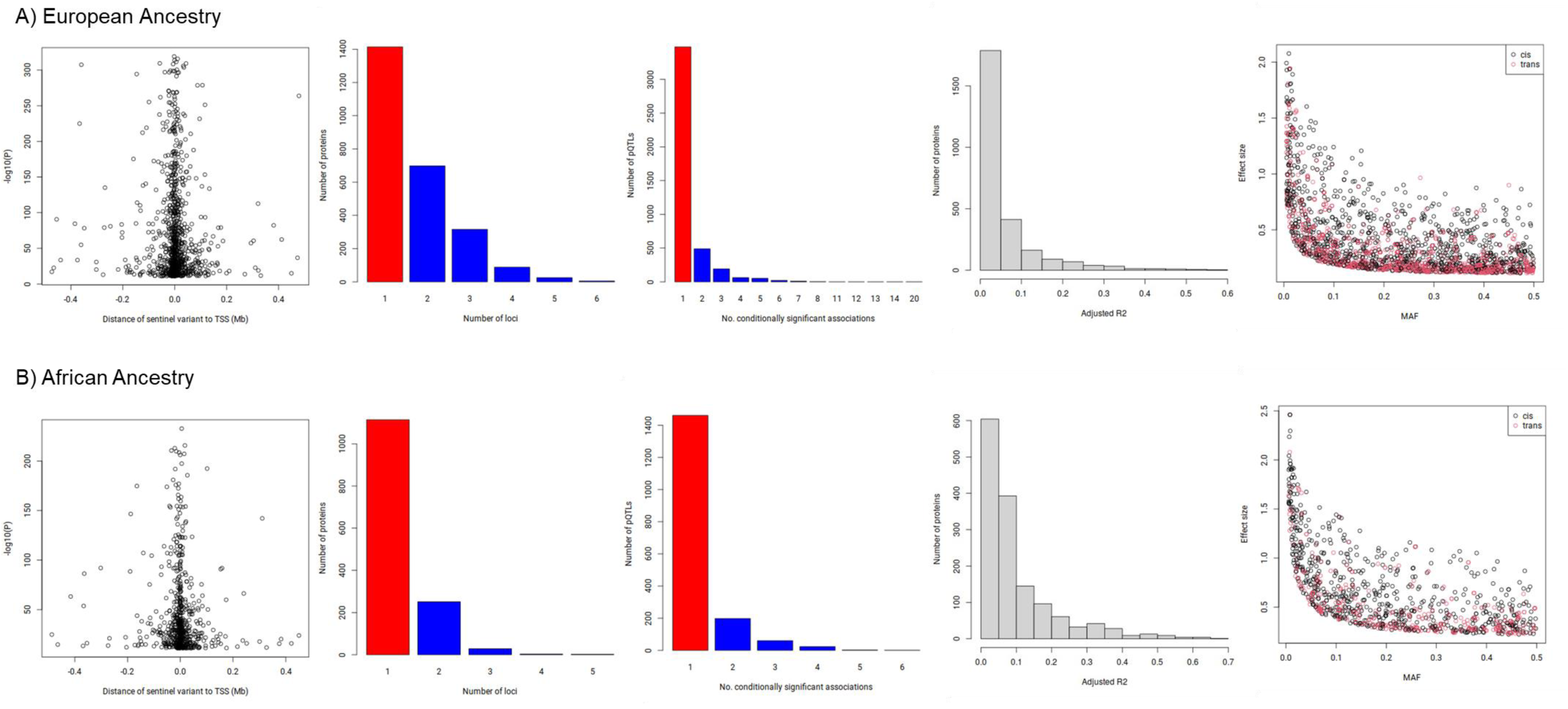
Visual summary of protein Quantitative Trait Loci (pQTLs) results by ancestry groups. From left to right: Upper Panel, significance of cis associations versus distance between sentinel variant to Transcription Starting Site (TSS); Number of significantly associated loci per aptamer; Number of conditionally significant associations within each pQTL; Bottom Panel, histogram of variance explained by conditionally significant variants; Effect size versus minor allele frequency.

In both AA and EA, most of the sentinel variants for cis pQTLs were located within 200kb (96%) and 100kb (90%/89%) respectively of the coding gene’s canonical transcription starting site (TSS), and 65%/58% respectively were within the gene itself. Most significantly associated proteins (56%, n=1,414) had a single pQTL, while 27% had two and 17% had 3-6 (**Figure 3**). In both EA and AA, the P-values of the cis-acting associations increased with distance of the sentinel variant to the TSS of the coding gene (**Figure 3**), mirroring previous pQTLs and eQTLs findings. (2,15) The majority of significantly associated proteins (AA: 80%, n=1,114 and EA: 56%, n=1,414) had a single pQTL, while 18% in AA (27% in EA) had two, and 2% in AA (17% in EA) had 3-6 respectively (**Figure 3**).We calculated the proportions of variance explained by the conditionally independent associated genetic variants for each of the 1,441 (AA) or 2,648 (EA) aptamers using GCTA (**Methods, Supplementary Tables 7** and **8**, **Figure 3**). The median variation in protein levels explained by pQTLs was 5.9% (25th percentile - 75th percentile: 3.4-12.6%) in AA and 2.7% (25th percentile - 75th percentile: 1.1-7.0%) in EA. For 197 (AA) / 187 (EA) proteins, genetic variants explained more than 20% of the variation. The maximum proportion of variance explained by a pQTL was 65% for Death-associated protein 1 (DAP) in AA with variant rs7614709, and 58% for Fc Fragment Of IgG Receptor IIIa (FCGR3A) in EA with variants rs17400517, rs79568124, rs4040189, rs546559546, and rs182782047. In both EA and AA, we observed a strong inverse relationship between effect size and MAF, consistent with previous GWAS of quantitative traits (**Figure 3**). In AA and EA, we found 38/102 and 151/331 associations (aptamer-sentinel variant pairs) with rare (MAF<1%) and low-frequency (MAF between 1-5%) variants respectively. Almost all of the strongest associations (per-allele effect size>1.5 SD) detected in both EA and AA involved rare or low frequency variants (51 among 52 associations in AA: 42 cis and 9 trans; 37 among 37 associations in EA: 20 cis and 17 trans).

### Shared pQTLs across EA and AA

We systematically compared the identified pQTLs across the two ancestry groups (**Methods**). Of 1,746 aptamer-region pairs pQTLs (involving 1,053 proteins) identified in AA, 1,113 (64% overall, 67% cis, 62% trans) were shared by EA, and 597 (involving 578 proteins, 34% overall, 31% cis, 36% trans) were distinct in AA (see **Methods** for definition of shared pQTLs), (**Supplementary Tables 3** and **9**). Of 4,315 aptamer-region pairs pQTLs identified in EA, in addition to the 1,113 shared pQTLs with AA (26% overall, 41% cis, 21% trans), and 2,481 (involving 2,168 proteins, 57% overall, 45% cis, 62% trans) were distinct in EA (see **Methods**), (**Supplementary Tables 4** and **10**).

Among the conditionally independent AA pQTLs (aptamer-variant pairs), 1,290 (60% overall, 58% cis, 60% trans) were identified in EA at the level of P<0.05/2,160=2.31×10^-5^ with the same variant or a variant at strong LD (R2>0.6) with a directionally concordant effect (**Supplementary Tables 5** and **11**). Among the conditionally independent EA pQTLs (aptamer-variant pairs), 2,306 (39% overall, 44% cis, 36% trans) were identified in AA at the level of P<0.05/5,901=8.47×10^-6^ with the same variant or a variant at strong LD (R2>0.6) with a directionally concordant effect (**Supplementary Tables 6** and **11**).

### Replication of previously reported pQTLs

We compared our pQTLs with previously reported pQTLs to demonstrate reproducibility and identify novel findings (**Methods**). Among 31,639 associated aptamer-region pairs derived from previously reported associations, 4,936 (15% overall, 40% cis, 13% trans) were replicated (P<1.58×10^-6^) in EA with the same variant or a variant at strong LD (R2>0.6); 1,493 (5% overall, 20% cis, 3% trans) were replicated (P<1.58×10^-6^) in AA; and 1,337 (4% overall, 17% cis, 3% trans) were replicated in both EA and AA (**Supplementary Table 12**). Of 1,746 aptamer-region pairs identified in AA, 808 (46% overall, 52% cis, 43% trans) were previously identified (**Supplementary Table 3**). For EA, of 4,315 aptamer-region pairs, 2,782 (64% overall, 75% cis, 61% trans) were previously reported (**Supplementary Table 4**). Interestingly, 53 out of 1,113 shared pQTLs across ancestries have not been previously reported (all in trans), advancing the knowledge on the genetic architecture of circulating proteome.

Among the 53 shared pQTLs across ancestries, 13 associations were observed with variant rs704 on chr17, nine with variant rs3733402 on chr4, and five with rs4976692 on chr5. The protein altering variant rs704 located in exon 7 of *VTN* was reported to be associated with blood levels of a high number of proteins.(1–3,13,16,17) The protein encoded by *VTN* functions in part as an adhesive glycoprotein. It is a lipid binding protein that forms a principal component of high-density lipoprotein. It can promote either cell adhesion or migration and is involved in a variety of other biological processes such as regulation of the coagulation pathway, wound healing, tumorigenesis and tissue remodeling. The protein altering variant rs3733402 located in exon 5 of *KLKB1* was reported to be associated with a high number of metabolites.(18–20). *KLKB1* encodes a glycoprotein that participates in the surface-dependent activation of blood coagulation, fibrinolysis, kinin generation and inflammation.

Among the 53 shared pQTLs across ancestries, we observed the largest effect for rs7649515 / rs2246888 with vascular endothelial growth factor D protein (VEGF-D). These variants reside in *MUSTN1* / *TMEM110-MUSTN1* and are in LD with missense variants in *ITIH4* and variants in *ITIH3*. ITIH3 has been recently reported as causal for HF risk.(21) VEGF-D is active in angiogenesis, lymphangiogenesis, and endothelial cell growth. We also identified the association of rs2246888 with VEGF-D. This variant has been reported associated with vascular endothelial growth factor D level in chronic kidney disease with hypertension and no diabetes.(17) We detected rs3733402 associated with VEGF sR3. This variant has been reported associated with vascular endothelial growth factor D levels and venous thromboembolism.(22,23) We detected rs12331618 associated with Low-density lipoprotein receptor class A domain-containing protein 4 (LDLRAD4). This variant has been reported associated with high-sensitivity cardiac troponin I concentration.(24) We reported the association of rs115655239 with L-lactate dehydrogenase A chain protein (LDHA). This variant was reported associated with serum levels of protein LDHB.(16)

### Functional and regulatory enrichment analysis

We accessed the enrichment of association analysis signals using GARFIELD in 1,005 features extracted from ENCODE, GENCODE and Roadmap Epigenomics projects, including genic annotations, chromatin states, histone modifications, DNaseI hypersensitive sites and transcription factor binding sites (**Methods**). In AA, among these 1,005 annotation features, 529 (53%) were found significantly (P<8.5×10^-5^) enriched (OR>1) and 55 (5%) depleted (OR<1) (**Supplementary Table 13**), with the most enriched feature being genic annotation: exon. The identified pQTLs were strongly enriched for variants in the coding regions (OR>2.5) and for locations in 3’ untranslated regions (UTR) (OR>2) (**Supplementary Fig. 1**). We also found relatively strong (OR>1.5, P<8.5×10^-5^) enrichment of pQTLs at features indicative of transcriptional activation in blood cells and at hepatocyte (chief functional cell of the liver) regulatory elements, as expected based on the role of the liver in protein synthesis and secretion (**Supplementary Fig. 1**). Similar findings were observed in EA, but with much more significantly enriched annotation features: 922 (99%) out of the 1,005 annotation features were found significantly (P<9.5×10^-5^) enriched (OR>1) and seven (<1%) depleted (OR<1) (**Supplementary Table 14**), with the most enriched feature being also genic annotation: exon. The identified pQTLs in EA were also strongly enriched for 5’ UTR (OR>2.5), in addition to variants in the coding regions (OR>4) and for locations in 3’ UTR (**Supplementary Fig. 1**).

### Assessment of Pleiotropy

We assessed the pleiotropy for all aptamers significantly associated with one or more variants (at the level of 5×10^-8^) and classified the pQTLs as protein-specific, vertical pleiotropic, or horizontal pleiotropic (**Methods**). Among the conditionally independent variants identified in AA (EA), 816 (1,576) were specific for a single protein, 106 (394) showed evidence of vertical pleiotropy, and 368 (1,266) showed evidence of horizontal pleiotropy (**Supplementary Tables 15 and 16**). An example of horizontal pleiotropy was rs3733402, a missense variant in the gene kallikrein B1 (*KLKB1*) on chromosome 4 (4q35.2), which showed significant association at the level of P<1.025×10^-11^ with 50 proteins in AA and 265 proteins in EA (**Supplementary Fig. 2**). For both AA and EA, the same GO term, 0007186 (G protein-coupled receptor signaling pathway), was shared by seven and 23 proteins, respectively, which were the maximum numbers of proteins sharing a single GO term. The SNP rs3733402 has been reported associated with various traits by GWASs including vascular endothelial growth factor D levels,(22) venous thromboembolism,(23) B-type natriuretic peptide levels,(25) serum metabolite levels,(19) and obesity-related traits.(26) An example of vertical pleiotropy in AA was a 5’ UTR variant (rs5471) located in gene *HP* that also mapped to the gene *TXNL4B* on chromosome 16 (16q22.2) was significantly associated with six proteins (**Supplementary Fig. 2**) and five out of these six proteins were related to the GO term “receptor-mediated endocytosis”. This variant was reported associated with LDL, HDL, and total cholesterol levels by GWASs.(27–29)

### Colocalization of pQTLs and eQTLs

To identify pQTLs for which the genetic associations are mediated by the effect of the variants at the transcriptional level, we compared the identified cis pQTLs with eQTLs derived from whole blood in GTEx (**Methods**). Among 1,050 cis pQTLs identified in EA after conditional analysis, 111 (11%) overlapped with an eQTL in the sense that the sentinel variant or a LD proxy (r2>0.8) was cis to the coding gene of the corresponding protein and was significantly (P<1.5×10^-11^) associated with the expression of this gene. For AA, 60 (9%) out of 643 cis pQTLs identified after conditional analysis overlapped with an eQTL. For each overlapped pair of pQTL and eQTL, colocalization test was performed over the region of the corresponding pQTL. Of the 111 overlapped pairs in EA, 64 (58%) had a highly (PP>0.5) or most highly likely (PP>0.8) colocalization, and thus shared a common causal variant (**Supplementary Table 4**). For AA, 25 out of the 60 (42%) overlapped pairs of pQTL and eQTL, colocalization was highly likely or most highly likely (**Supplementary Tables 3**). **Supplementary Table 17** lists the 14 pairs of pQTLs and eQTLs found to highly likely colocalize in both EA and AA. For seven of these pairs, the candidate causal variant was identical for EA and AA. For the other pairs, the candidate variants in EA and AA were in high LD (r2≥0.85). Several of the causal variants identified are in high LD with a missense variant in the coding gene of the target protein (rs291466 - *HIBCH*, rs12480408 - *CPNE1*, rs73920010 - *TP53I3*, and rs4721057 - *TMEM106B*).

### Colocalization of pQTLs and disease-related traits

We assessed colocalization between the pQTLs identified in ARIC AA and EA samples and recently reported GWAS signals for different disease-related traits (**Methods**). Among pQTLs identified in EA and AA, 183 vs 49 proteins respectively were found to colocalize with one or more traits, and 61 vs 14 potentially causal variants were identified. A total of 371 signals highly likely to colocalize were identified (8% in cis), involving 191 proteins and 65 causal variants, among which 138 (31%) were shared across EA and AA (**Supplementary Tables 18** and **19**). The 65 causal variants are located on 16 chromosomes and can be grouped into 29 meta-regions (**Figure 4**). Among all 196 mega region-trait-protein triplets, 140 had just one candidate causal variant, and the rest of them had 2∼4 different (but adjacent) candidate causal variants obtained for samples of different ancestries (EA/AA), different studies or different sub-phenotypes. For HF, we identified 124 colocalized proteins in 10 meta-regions on eight chromosomes and there were 36 candidate causal variants (**Supplementary Fig. 3**). Previous studies highlighted the relevance of some of these proteins for HF such as IL-23,(30,31) VCAM-1,(32) SEM4D,(33) Spondin-1,(34) VEGF sR2 and sR3 (35,36), sE-Selectin,(37,38) IGF-I sR,(39) Endoglin,(40) and GLCE.(41) Proteomic MR causally implicated proteins such as GLCE and Spondin-1 with HF risk.(41,42)

**Figure 4.**
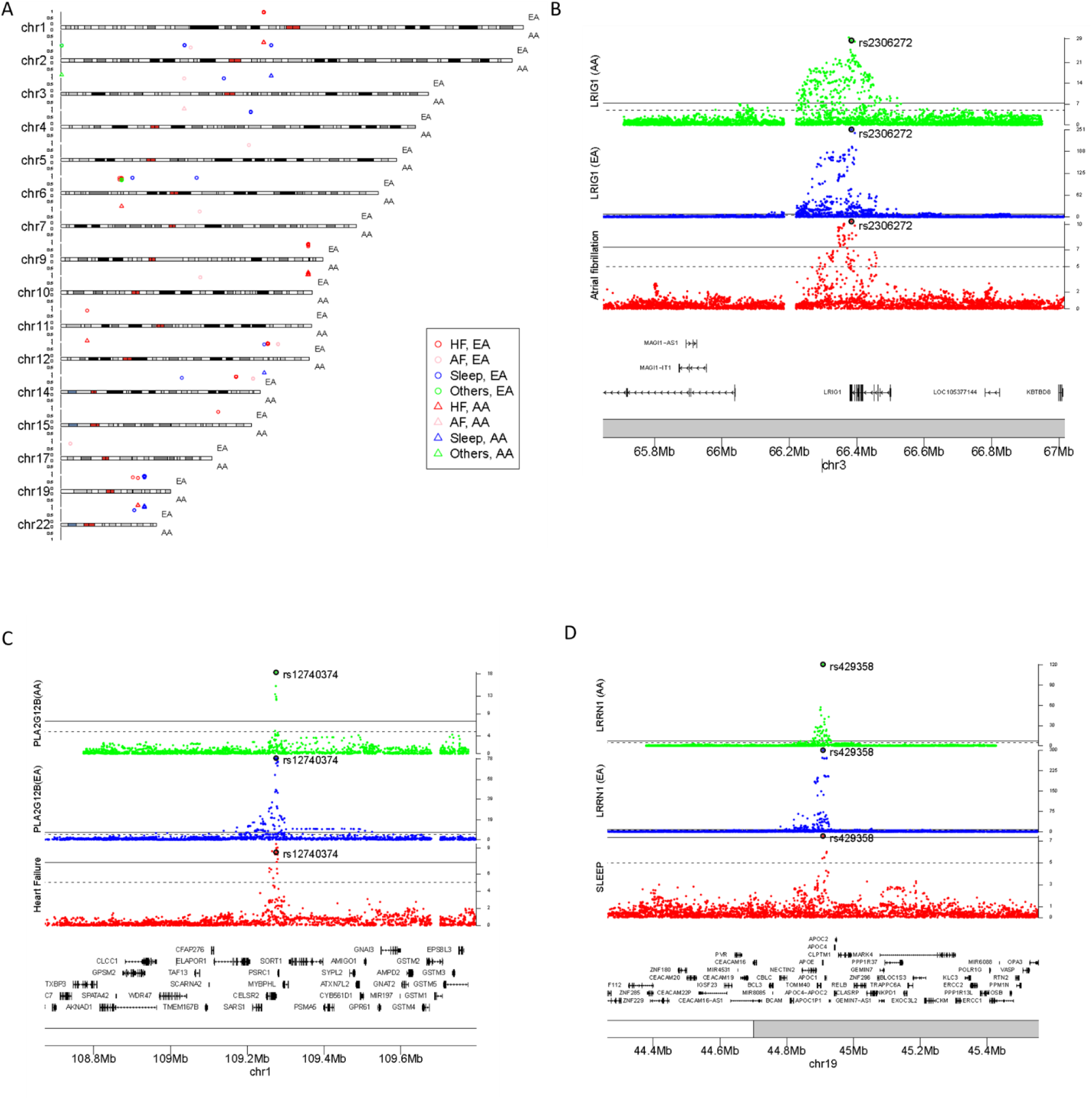
Colocalization of protein Quantitative Trait Loci (pQTLs) and genome-wide association signals of phenotypes. (A) Posterior probability of colocalizations of all four types of traits (Heart Failure (HF), Atrial Fibrillation (AF), sleep and others including fractional anisotropy, retinal vascular density, and retinal vascular fractal dimension) with pQTLs identified from European (EA) and African (AA) ancestries. (B) Significant associations of protein levels of LRIG1 in EA and AA and AF (3) in a region around the identified common causal variant, rs2306272, a missense variant in LRIG1. (C) Significant associations of protein levels of sPLA(2)-XIII encoded by *PLA2G12B* in EA and AA and HF (90) in a region around the identified common causal variant, rs12740374, a downstream gene variant at *PSRC1* and 3’ UTR variant at *CELSR2*. (D) Significant associations of protein levels of LRRN1 in EA and AA and sleep (84) in a region around the identified common causal variant, rs429358, a missense variant at *APOE*.

In total, there were 16 unique protein-trait of CVD type (HF or AF) pairs for which we identified colocalization signals for both EA and AA with the same candidate causal variants (**Supplementary Table 18**). These associations were in trans, and we also observed some pleiotropy with for example rs947073006 associated with six proteins and rs879055593 associated with four proteins. For example, for both AA and EA, HF and sPLA(2)-XIII (coded by *PLA2G12B*) were found to colocalize in the locus 1p13 with the same causal variant, rs12740374 (classified as horizontal pleiotropic), a downstream gene variant at *PSRC1* and 3’ UTR variant at *CELSR2* (**Figure 4**). Another example includes atrial fibrillation (AF) and LRIG1 that were found to colocalize in the locus 3p14 with the same causal variant, rs2306272 (classified as protein specific), an intron variant in *SLC25A26* and a missense variant in *LRIG1* (**Figure 4**). We also observed shared colocalization signals with sleep between EA and AA with the same candidate causal variants for 18 proteins (**Supplementary Table 18**). These associations were in trans, and we also observed important pleiotropy with the variant rs429358 (classified as horizontal pleiotropic) defining *APOE* haplotype, the major genetic risk factor for late-onset Alzheimer’s disease, reported for most of the colocalizations. Sleep and LRRN1 colocalized in 19q13.3 with the same causal variant rs429358 (**Figure 4**).

### Phenome-proteome-wide Mendelian randomization (MR) analyses

Utilizing the protein-associated variants identified in EA and shared and/or validated in AA as genetic instruments, we conducted MR analyses to identify plasma proteins with potential causal effects on various outcomes (**Methods**). After LD-clumping, we retained 992 aptamers (representing 971 unique targeted proteins) as exposures, each of which had one or more conditionally independent instruments from a set of 1,045 variants. We used these instruments to conduct two-sample MR to evaluate evidence for causal effects on 242 phenotypes (16 diseases and 226 intermediate traits). At a Bonferroni-corrected threshold of P<3.62×10^-7^, we identified 565 protein-phenotype associations with MR evidence (**Supplementary Table 20, Supplementary Fig. 4**, involving 252 aptamers (representing 245 targeted proteins) and 63 phenotypes (18 diseases and 48 intermediate traits). A total of 28 associations were detected for five disease categories, among which nine were previously reported by phenome-wide proteomic MR. Among the 565 associations with MR evidence, 58 were previously reported by phenome-wide proteomic MR, involving 43 aptamers (39 targeted proteins) and 24 phenotypes (five diseases and 19 intermediate traits). In the rest of the 507 new associations, 19 represented a candidate causal effect of 14 proteins on 14 diseases (**Supplementary Table 21**). Among these 19 MR-based associations, we found that seven have been reported by disease-specific proteomic MR study such as MOTI vs. type 2 diabetes, (43) TFF3 vs. rheumatoid arthritis, (44) or PYY vs colorectal cancer (45) (**Supplementary Table 21**). To our best knowledge, the remaining 12 associations seem to represent potential causal relationships that have not been previously reported (**Figure 5**). The top associations for each disease category were: Granzyme B vs. primary sclerosing cholangitis, TFF3 vs. ulcerative colitis, TFF3 vs. malignant non-melanoma skin cancer, and BIN1 vs. Alzheimer’s disease.

**Figure 5.**
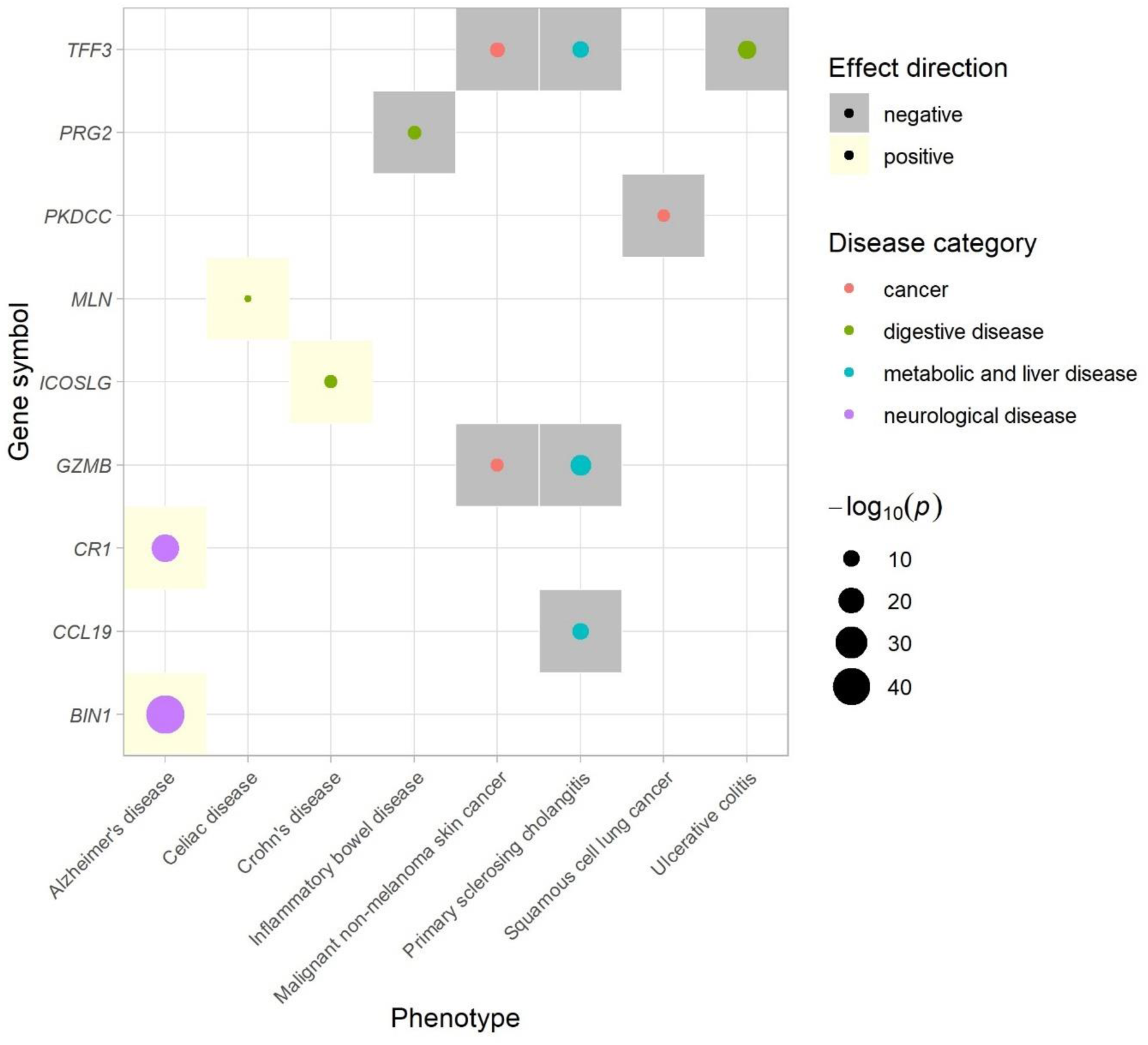
Mendelian Randomization (MR) analyses of plasma proteins’ effects on human diseases. The plot shows 12 statistically significant (P<3.62×10^-7^) Mendelian Randomization (MR) findings of selected human diseases, with each protein-disease pair being represented by a filled circle. The diseases were grouped into five categories (cancer, digestive, immunological, metabolic, and liver, and neurological) and are represented by different colors. The sizes of the circles emphasize the strength of the association. The colors of background square boxes represent the direction of effect. The x-axis corresponds to the diseases listed in alphabetical order and the y-axis corresponds to the gene symbol of the proteins.

## Discussion

By analyzing up to 9,455 ARIC participants representing two ancestries (7,584 EA and 1,871 AA), we have identified 1,746 and 4,315 significant associations between regional sentinel variants and proteins in AA and EA participants, respectively. By analyzing both cis and trans genetic variants, we increased the number of pQTLs previously described in the ARIC study. (46) The mean variation in protein levels explained by the pQTLs (0.10 in AA and 0.06 in EA) was consistent with published pQTL studies.(8,46) The majority of the pQTL associations were shared across EA and AA (>60%), and a higher proportion of shared cis-QTLs were colocalized with eQTLs compared to cis-QTLs not shared across EA and AA. We also observed that many of the novel pQTLs, particularly the trans-pQTLs, were identified in AA participants (57% of trans-associations identified in AA not previously reported vs 39% in EA). By utilizing the pQTLs generated, multiple plausible causal pathways were identified, providing insights into disease molecular mechanisms.

By leveraging EA and AA, our analysis shed light on potential ancestry-specific pQTLs (34% of AA associations, and 57% of EA associations). We identified five AA-specific trans-pQTLs (**Supplementary Table 22**) that did not overlap with the region of EA pQTLs with the same protein. None of these pQTLs were previously reported by studies conducted in participants of European ancestry. We identified 640 EA-specific pQTLs (**Supplementary Table 23**) that did not overlap with the region of AA pQTLs with the same protein or any AA-pQTLs reported in Katz et al. At least for some of ancestry-specific pQTLs, the difference in the association of the protein level and the genomic locus between ancestries may reflect a complex interplay between a pQTL and other genetic variants, which has been previously observed in the gene expression analysis,(47) and/or environmental factors that could be investigated in future studies.

The identified pQTLs provided us with an opportunity of conducting both a proteome-wide colocalization screen for GWAS traits and a phenome-proteome wide, hypothesis-free scan of potential causal effects of proteins on diseases and other complex traits. We identified a total of 371 colocalization signals, mainly in trans, involving 191 proteins and 65 potentially causal variants, and demonstrated the utility of pQTLs in pinpointing potential causal genes and genetic variants from a plethora of GWAS signals. The identification of colocalization signals shared by data from different ancestries provided even stronger evidence of a causal genomic variant. A cis example included AF and LRIG1 colocalizing in 3p14 with the candidate causal missense variant rs2306272 in *LRIG1* in EA and AA. A recent study identified LRIG1 as promising AF drug target with genetically predicted protein LRIG1 decreasing AF risk. (48,49) Leucine-rich repeats and immunoglobulin-like domains (LRIG) proteins play a role in the regulation of lipid metabolism, and common genetic variants in LRIG1 have been described associated with an increased risk of BMI but a reduced risk of T2D, a risk factor for AF.(50)

As trans-pQTLs are more likely to be pleiotropic than cis-pQTLs, we carefully evaluated pleiotropy of the genetic instruments selected for MR. We identified 565 protein-phenotype associations with MR evidence involving 250 aptamers and 61 phenotypes, and illustrated how the pQTLs of vertical pleiotropy identified across ancestries can be leveraged to identify potential causal factors or drug targets for complex diseases and distinguish those causal factors from potential biomarkers. Well known examples include the Agouti-signaling protein ASIP with melanoma skin cancer, PCSK9 with LDL-C and total cholesterol, PAPP-A with IBD, and angiotensinogen with diastolic and systolic blood pressure. (5,7) We identified 12 potentially novel causal associations (total of six variants in cis and four variants in trans) between 10 proteins and nine diseases, including: Granzyme B with primary sclerosing cholangitis, TFF3 with malignant non-melanoma skin cancer, EMBP with inflammatory bowel disease, PKDCC with squamous cell lung cancer, and BIN1 with Alzheimer’s disease.

Recently, Granzyme B has been shown to induce apoptosis and fibrosis in sclerosing cholangitis.(51) TFF3 has been shown to be significantly upregulated in cancer (such as gastric cancer, colorectal cancer, lung cancer, thyroid cancer, and breast cancer) and to play a key role in tumor progression and metastasis.(52,53) Due to its role in cancer progression and metastasis, TFF3 is a potential biomarker and therapeutic target for cancer. Bone marrow proteoglycan (EMBP/PRG2) was identified as a potential biomarker for active Crohn’s disease.(54–56) PKDCC has been suggested to act as an oncogene in certain cancer types due to its role in cellular signaling pathways. Indeed, protein kinases are key regulators of cellular processes such as growth, differentiation, and apoptosis, which are often dysregulated in cancer cells. PKDCC regulates Wnt/PCP signaling and Wnt signaling is involved in the regulation of proliferation and migration of various tumor cells.(57) Out of the 10 proteins, PKDCC is a potential drug target (Human Protein Atlas). *BIN1* is the second most important genetic risk factor for late onset Alzheimer’s disease (AD) and is associated with amyloidopathy and tauopathy. BIN1 interacts with tau, one hallmark protein in AD and affects its aggregation and spreading within the brain.(58,59) BIN1 also influences the processing and clearance of amyloid beta, another hallmark protein in AD.(60,61) However, to date, no consensus has been reached on the role of BIN1 in AD pathogenesis.(62–64)

Strengths of this study include the large number of plasma proteins analyzed in a relatively large sample of diverse participants from a well-characterized cohort. Compared to the great proportion (60-66%) of pQTLs identified in AA participants that were validated in EA participants, consistent with other pQTL studies, (8) a smaller proportion (39-43%) of pQTLs identified in EA participants were validated in AA participants. Reasons for not validating some of the pQTLs identified in one ancestry in the other ancestry include differences in sample sizes and thus power to detect associations, LD-patterns, and allele frequencies. We detailed a strategy to assess the sharedness of the pQTLs identified across ancestries using more stringent threshold than for validation. It is important to note that because of the regional nature of the pQTLs and the existence of LD between variants that potentially differs between ancestries, a pQTL may have different sentinel variant and/or region’s coordinates in different ancestry groups, although the regions should largely overlap, making the determination of sharedness of pQTLs between AA and EA nontrivial. Our classification of MR associations as previously reported relied on phenome-wide proteomic MR studies and complemented the classification with a lookup of potentially novel causal associations in disease-specific MR studies. Finally, we have not explored the effects of very rare variants on plasma proteins. Future work should focus on evaluating the impact of such variation on the heritability of plasma proteins.

## Conclusion

Our study demonstrates that an important proportion of pQTLs is distinct across European and African ancestries and provides novel insights into the genetic regulation of plasma protein levels, specifically by distal variants, in these two groups. More research is warranted on the trans pQTLs identified in this study, particularly the 53 shared pQTLs not previously reported and co-identified in both European and African ancestries. Further studies aiming to disentangle the causes for discordant pQTLs between European and African ancestries should be conducted to improve our understanding of both the genetic architecture of proteome and the multi-ancestry differences of circulating protein levels and evaluate their implications in different etiologies of complex diseases across populations.

## Methods

### Study populations

We leveraged participants from the Atherosclerosis Risk in Communities (ARIC) Study, an ongoing community-based cohort study of 15,792 participants that initially enrolled during 1987 and 1989 from four communities across the US: Washington County, Maryland; suburbs of Minneapolis, Minnesota; Forsyth County, North Carolina; and Jackson, Mississippi. (8,65) The third visit (v3) occurred in 1993-1995, when blood samples used for the measurement of the proteome were collected for the current study. The ARIC study was approved by the institutional review boards at each ARIC center, and participants provided informed consent.

### Proteomic measurements

The relative concentrations of plasma proteins or protein complexes from the blood samples were measured by SomaLogic Inc. (Boulder, Colorado, US) using an aptamer (SOMAmer)-based approach, (66) which selected aptamers for almost any protein target efficiently. A total of 5,284 aptamers were measured, and we excluded aptamers that had coefficient of variation > 50%, variance < 0.01, or binding to mouse Fc-fusion, a contaminant, or non-proteins. As a result, 4,877 aptamers measuring 4,651 unique proteins or protein complexes were included in the current analyses. Quality control of the ARIC Study proteome data has been described in detail previously. (67)

### Genotyping and imputation

Genotype data for ARIC was completed using Affymetrix Array 6.0. Quality metrics used to filter variants for the imputation basis and association testing included sex mismatch, discordance with previously genotyped markers, first-degree relative of an included individual, and genetic outlier based on allele sharing and principal components analyses, insufficient call rate (> 5%), and deviation from Hardy-Weinberg equilibrium (HWE, P<1×10^-5^). Imputation was performed on the data passed quality filters in two steps: 1) Pre-phasing with SHAPEIT (v1.r532), and 2) Imputation using the Trans-Omics for Precision Medicine (TOPMed) reference panel (Freeze 5b, build 38). (68) A total of 18,098,426 (AA) and 9,681,469 (EA) variants that had Rsq ≥ 0.3, MAF ≥ 0.5% and HWE P > 5×10^-6^ were considered in the current analyses. After excluding participants who did not have cleaned plasma protein and/or genotype data, 1,871 AA and 7,584 EA participants were retained in the current study.

### GWAS and functional annotation of variants

For each aptamer, a linear regression was first applied to the log2-transformed protein levels adjusting for age, sex, and study center in each ancestry. The residuals from the linear regression were rank-inverse normalized. The resulting phenotype was then used for genome wide association testing of autosomal SNPs with MAF ≥ 0.5%. Association tests were conducted by ancestry using linear regression on allelic dosages using RVTESTS v20190205, (69) adjusting for age, sex, study center, and the first three principal components of ancestry to account for population stratification. Genetic associations were considered genome-wide and proteome-wide significant if P<1.025×10^-11^ (GWAS threshold of 5×10^-8^ corrected for 4,877 aptamers). Functional annotations were performed on the SNPs that were detected significantly associated with one or more aptamers using the Ensembl Variant Effect Predictor (VEP). (70) All functional consequences of a variant or a gene were listed, whether the gene is the coding gene of the target protein of the associated aptamer or not.

### Refinement of pQTLs

Many of the adjacent variants associated with an aptamer may represent one single signal due to LD. To establish a list of conditionally independent loci significantly associated with any given aptamer and facilitate comparison with published findings, we used the following approach. (2) First, we defined 1-Mb regions around each significant variant for a given aptamer and merged any overlapping regions. In the resulting regions of each aptamer, we defined a regional sentinel variant as the variant with the smallest P-value in the region. Because of the complexity of the Major Histocompatibility Complex (MHC) region, we treated it as a single region: Chr6: 25,499,772-34,032,223 (Build 38). For regions associated with different aptamers, a LD-based merging approach was used to combine regions if their regional sentinel variants were the same or in high LD (r2 ≥ 0.8). A cis-pQTL was defined as a sentinel variant within 1MB to the TSS of the coding gene of the target protein of the associated aptamer. A trans-pQTL was defined as a sentinel variant outside ±5MB of the TSS of the coding gene of the target protein of the associated aptamer or located on another chromosome.

We then used GCTA v1.93.2(71,72) with ARIC genotypes as LD reference panel to perform a stepwise model selection procedure to select conditionally independent variants in each region for a given aptamer. A single COJO analysis using the “cojo-slct” option was carried out for each aptamer. To properly calculate the proportion of variance explained by each variant in the model selection procedure, GWAS summary data for all 18,098,426 and 9,681,469 variants after filtering with P_HWE_ > 5×10^-6^ were used as input for GCTA for AA and EA respectively. To partially reduce computational burden, the genotypes for all significant variants (P<1.025×10^-11^) in AA or EA were used to estimate pairwise LD. Therefore, occasionally, a variant that was not significantly associated with an aptamer in the single variant association test might appear in the list of conditionally independent associated variants in the COJO analysis, and in a few instances, the regional sentinel variant was not included in the list of conditionally independent associated variants. (2) In those cases, we conducted a joint analysis with GCTA using the “cojo-joint” option to estimate the joint effects of the sentinel variants and those identified in the stepwise model selection procedure.

### Comparison between EA and AA pQTLs

We evaluated the cross-ancestry validation of the identified pQTLs. For each pair of aptamer and conditionally independent associated variants obtained for AA (EA), if the variant or a proxy (r2 > 0.6, in the ancestry of AA (EA)) was found to be significantly associated with the same aptamer in EA (AA) at a Bonferroni-corrected P-value (P<0.05/2,160=2.31×10^-5^ for AA; P<0.05/5,901=8.47×10^-6^ for EA), and had a concordant direction of effect, this pQTL was considered to be validated. We conducted the same type of cross-ancestry validation for pQTLs represented by pairs of aptamer and regional sentinel variants. The P-value thresholds for validation were P<0.05/1,746= 2.86×10^-5^ and P<0.05/4,315=1.16×10^-5^ for AA and EA respectively.

To determine whether a pQTL was shared between EA and AA at the region level, we identified pQTLs that were validated in the other ancestry at a more stringent level of P<1.025×10^-11^. For example, if a pQTL (i.e. an aptamer-region pair) identified in AA (EA) (referred as an AA (EA) pQTL) was validated in EA (AA) at the level of P<1.025×10^-11^ by the sentinel variant or its proxy (referred to as validating variant), the AA (EA) pQTL and the corresponding EA (AA) pQTL are said to be the same pQTL shared by EA and AA. An AA pQTL that was validated at P<1.025×10^-11^ in EA did not necessarily mean that an EA pQTL was also validated at P<1.025×10^-11^ in AA. Indeed, if the validating variant of the AA pQTL (which must reach P<1.025×10^-11^ in EA) was not the proxy of the EA sentinel, the sentinel variant of the EA pQTL (or its proxies) might not have reached the P<1.025×10^-11^ level in AA.

In the case where no EA (AA) pQTL was validated in AA (EA) at P<1.025×10^-11^ via a variant that could be found in the region of the AA (EA) pQTL and it was not validated in EA (AA) either, the AA (EA) pQTL was not a shared pQTL with EA (AA). If the validation level did not even reach the nominal level P<0.05, it was considered as a candidate of AA (EA)-specific pQTLs.

### Replication of existing pQTLs

We attempt to collect all pQTLs previously identified in the literature and assess whether they replicated by our data. We first performed a literature search using the NCBI Entrez utility in R (rentrez) for pQTL studies published from 2017 onward (3,4,8,9,12,13,46,73–76) and merged the resulted list of pQTLs with the list given by Sun et al., which represented the pQTLs discovered between 2008 and 2016. (2) Studies were included only when plasma or serum samples were profiled. Only data for proteins also assayed and included in our GWAS were included. Then associations with P<1.025×10^-11^ were clumped into separate regions in the same way as for our data. For each of the resulting regions and each of the associated aptamers, we checked whether its sentinel variant or a proxy with r2 > 0.6 was associated with the same aptamer/protein in our study at a given significance threshold (P<0.05/31,639=1.58×10^-6^). Lifting over to hg19 was performed to our data prior to the replication analysis of previously reported pQTLs.

### Functional and regulatory enrichment analysis

To test whether the pQTLs identified from our GWAS were enriched for functional and regulatory elements, we applied GARFIELD (77) to the summary statistics data obtained for all aptamers significantly associated with a variant. GARFIELD v2 is a tool to assess the enrichment of association analysis signals in 1,005 features extracted from ENCODE, GENCODE and Roadmap Epigenomics projects, including genic annotations, chromatin states, histone modifications, DNaseI hypersensitive sites and transcription factor binding sites, among others. It first performs greedy pruning of variants (LD r2 > 0.1) and annotates them based on functional information overlap with the 1,005 annotation features. Then, it quantifies enrichment using odds ratios (OR) at various GWAS p-value cutoffs and assesses their significance by employing generalized linear model testing, while accounting for confounding factors, such as minor allele frequency, distance to nearest transcription starting site and number of LD proxies (r2 > 0.8). We performed the enrichment analysis for all variants tested in the GWAS and used the significance level P=1.025×10^-11^ as the cutoff. For AA, after the pruning process, 8,776,031 conditionally independent variants were selected for enrichment analysis. The effective number of annotations was estimated as Meff=588 and the enrichment p-value adjusted for multiple testing was set to P_adj_=8.5×10^-5^. For EA, after the pruning process, 756558 conditionally independent variants were selected for enrichment analysis. The effective number of annotations was estimated as Meff=528 and the enrichment p-value adjusted for multiple testing was set to P_adj_=9.5×10^-5^.

### Assessment of Pleiotropy

Proteins can be correlated due to different mechanisms. Vertical pleiotropy refers to proteins that act in a cascade-like manner and are associated within a pathway. On the other hand, horizontal pleiotropy implies that proteins are acting through different pathways. Classification of these two types of pleiotropy is of importance in downstream analysis, such as Mendelian Randomization (MR) methods. We assessed the pleiotropy of the pQTLs using a method like the approach proposed in Pietzner et al. (78) For all the aptamers significantly associated with one or more variants at the level of 5×10^-8^, all GO-terms referring to biological processes were retrieved from the UniProt database using the UniProt-IDs as query. Then each of the variants with one or more significantly associated aptamers was categorized as follows: (1) Solely associated with a specific protein (Type 1); (2) All associated aptamers belonging to a single GO-term (Type 2); The majority (>50%) belonging to a single GO-term (Type 3); and No single GO-term covering more than 50% of those aptamers (Type 4). Type 1 represents a protein-specific association, Types 2 and 3 are referred to as vertical pleiotropy, and Type 4 as horizontal pleiotropy.

### Colocalization Analyses

We leveraged expression QTL (eQTL) data from whole blood from The Genotype-Tissue Expression (GTEx) project. (79) A pQTL was considered overlapping with an eQTL if the sentinel variant or any of its LD proxies (r2 > 0.8) was lying in cis of (i.e., within 1Mb) the coding gene of the pQTL protein and was significantly (P < 1.5×10^-11^) associated with the expression level of this gene. The LD proxies of the sentinel variants were queried using the R package LDlinkR using an appropriate reference population from the 1000 Genomes Project (that is, CEU and ASW for EA and AA, respectively). Then, for each of the cis pQTLs with an overlapping eQTL, colocalization was performed in the region of the pQTL using the Coloc package. (80) Evidence of colocalization between a pQTL and an eQTL was assessed using the posterior probability (PP) of colocalization. If PP > 0.5, the association was deemed as “likely to colocalize”, and when PP > 0.8, the pQTL and eQTL pair was deemed as “highly likely to colocalize”.

To evaluate colocalization of pQTLs identified in ARIC AA and EA samples and disease/trait associations, the testing regions using Coloc were chosen as the regions of the pQTLs in which there were significant (P < 5×10^-8^) association signals for the diseases/traits of consideration. GWAS summary statistics data were downloaded from 12 studies on the following four types of diseases/traits: cardiovascular disease (CVD) heart failure (HF) and atrial fibrillation (AF), sleep measures, and others including fractional anisotropy, retinal vascular density, and retinal vascular fractal dimension. (81–92) These GWASs were mainly performed in European populations (N=8). A few studies were performed in multi-populations (N=3 including ECG traits, stroke, and AF) and one was performed in African populations (stroke). Default priors were used and as described above, associations with PP4 (posterior probability of colocalization) > 0.5 were deemed “likely to colocalize” and PP4 > 0.8 “highly likely to colocalize”.

### MR analysis

A phenome-proteome-wide two-sample MR mapping was performed to infer causal effects of plasma proteins on hundreds of phenotypes and traits, for which GWAS summary statistics are publicly available. We used GWAS summary statistics data that was collected and curated manually for MR-Base (93) including 145 outcomes (phenotypes and traits) from the first batch and 97 phenotypes from the second batch (ieu-a and ieu-b). As most (if not all) of these summary statistics were drawn from studies for populations of European ancestry, we selected exposures and instrumental variables (IVs) for MR analysis from the list of conditionally independent associated pairs of aptamer and variants identified in EA (**Supplementary Table 7**). A pair of such aptamer and variant was chosen for MR analysis if (a) the pQTL defined by the pair of the corresponding genomic region and aptamer in EA was shared and/or validated in AA (see above for the definitions of shared and validated pQTLs); and (b) the variant was classified as either protein-specific or vertical pleiotropic. For each exposure (protein), the IVs were clumped at r2 < 0.001. Wald test was used if there was only a single IV available for a pair of outcome and exposure, and inverse weighted variance tests were used if at least two SNPs were analyzed together. Sensitivity tests included Cochran’s heterogeneity and MR-Egger intercept test of pleiotropy. Unique pairs of exposure and outcome were obtained by removing duplicated phenotypes, keeping the one with largest sample size. SNPs detected as pleiotropic were evaluated at 4.29×10^-5^ (0.05/1,165 total number of unique pairs of exposure and outcome) to ensure the validity of the instruments used. To correct for multiple testing, the causal relationship of a protein on a phenotype was defined as statistically significant if the corresponding MR P-value was lower than 3.62×10^-7^ (0.05/138,103 total number of tests performed). Finally, we compared our significant MR results with published proteomic MR studies (5–7) to categorize our findings as novel or previously reported. All analyses were performed using “TwoSampleMR” package (version 0.5.6) in R. (94)

A flowchart detailing our analysis strategy is presented in **Figure 1**.

## Supporting information

Supplementary Figures and Small Supplementary Tables

Large Supplementary Tables

## Authors contributions

C.S and J.M contributed equally to the study. B.Y and N.C conceived the project; J.M, N.Q.H.N carried out all analyses with supervision from C.S and B.Y; C.S and J.M drafted the manuscript; B.Y, E.B, J.C, R.C.H, C.M.B, A.M, and N.C provided comments. All authors reviewed and approved the final version of the manuscript.

## Acknowledgements and Funding

The Atherosclerosis Risk in Communities study has been funded in whole or in part with Federal funds from the National Heart, Lung, and Blood Institute, National Institutes of Health, Department of Health and Human Services, under Contract nos. (75N92022D00001, 75N92022D00002, 75N92022D00003, 75N92022D00004, 75N92022D00005). The authors thank the staff and participants of the ARIC study for their important contributions.

SomaLogic Inc. conducted the SomaScan assays in exchange for use of ARIC data. This work was supported in part by NIH/NHLBI grant R01 HL134320 and R01 HL148218. Dr. Sarnowski is funded through NIA R00AG066849. Dr. Yu is funded through NHLBI R01 HL148218. Dr. Ballantyne is funded through NHBLI R01 HL134320.

## Data availability

The phenotypic and genotypic data used from the ARIC Cohort can be accessed via dbGaP (Study Accession: phs000280.v8.p2) or BioLINCC (https://biolincc.nhlbi.nih.gov/studies/aric/). The ARIC proteomic data can be requested through the study’s data coordinating center upon an approved manuscript proposal and Data and Materials Distribution Agreement. Genome-wide summary-level statistics for single-SNP pQTL analysis can be accessed at https://www.synapse.org/Synapse:syn63141185.

