## Supplementary Figures and Small Supplementary Tables for "Ancestrally diverse genome-wide association analysis highlights ancestry-specific differences in genetic regulation of plasma protein levels"

**Sarnowski C, Ma J et al**

### Contents

**Supplementary Figures** .......... 2

**Supplementary Figure 1.** GARFIELD functional and regulatory enrichment analysis. .......... 2

**Supplementary Figure 2.** Chord plots showing association of a genetic variant with proteins by ancestry groups for two examples of pQTLs classified as horizontal or vertical pleiotropic 10

#### Supplementary Figures

#### **Supplementary Figure 1.** GARFIELD functional and regulatory enrichment analysis.

A) European ancestry

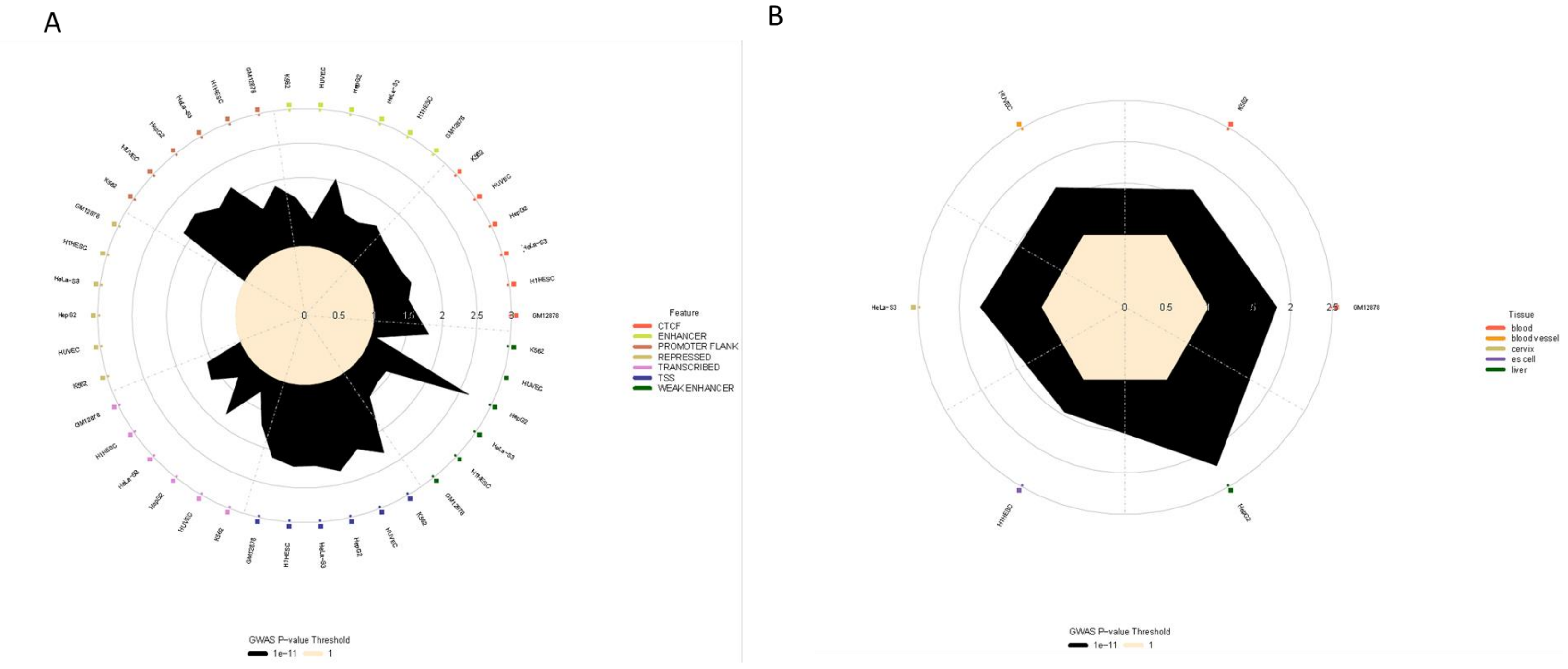

Chromatin States FAIRE

C

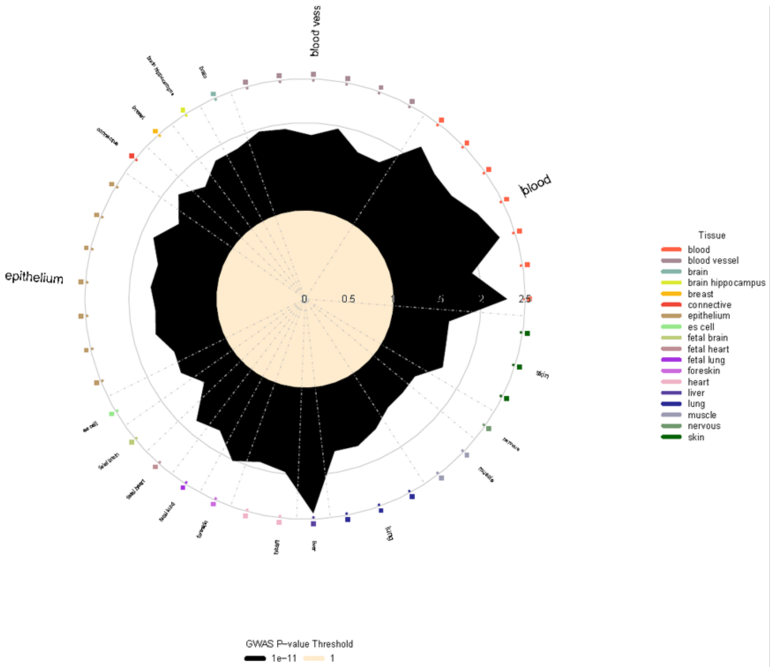

Footprints

D

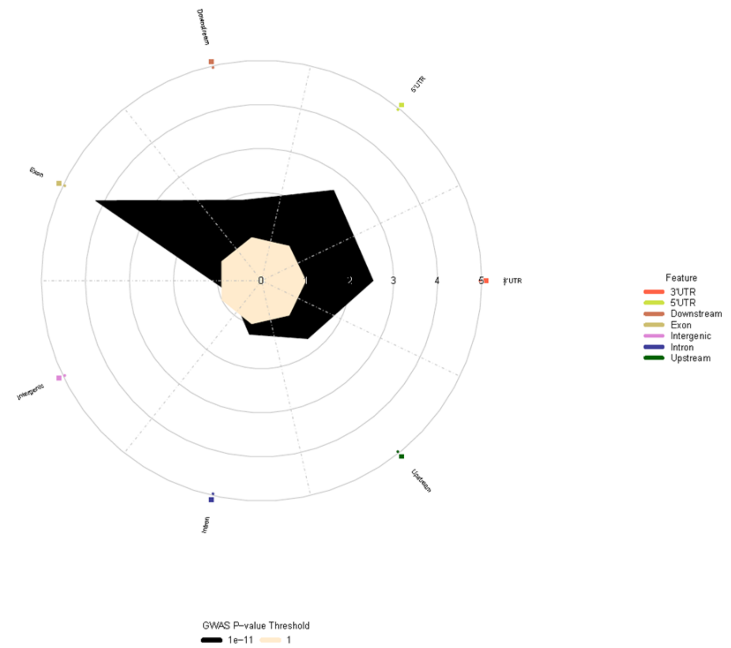

Genic

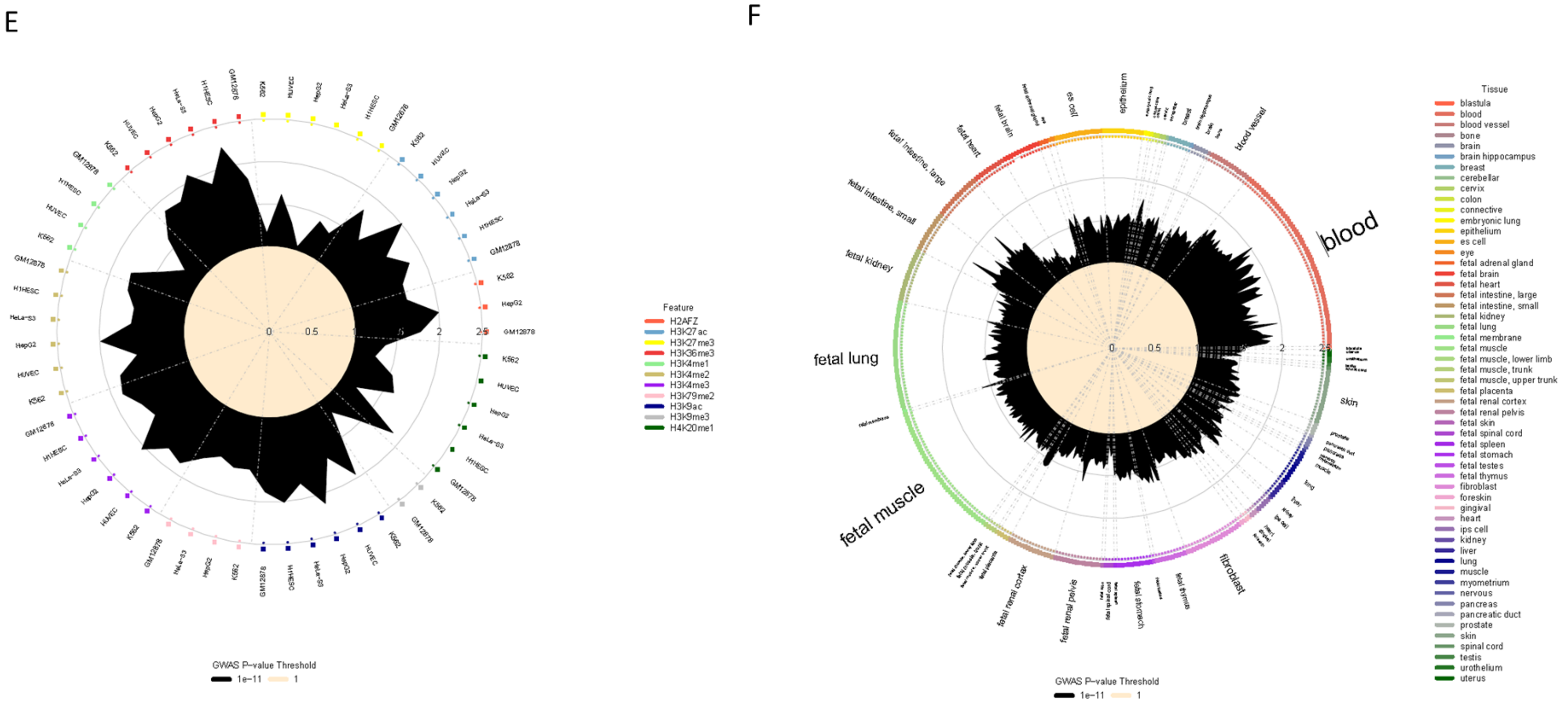

Histone Modifications

Hotspots

G

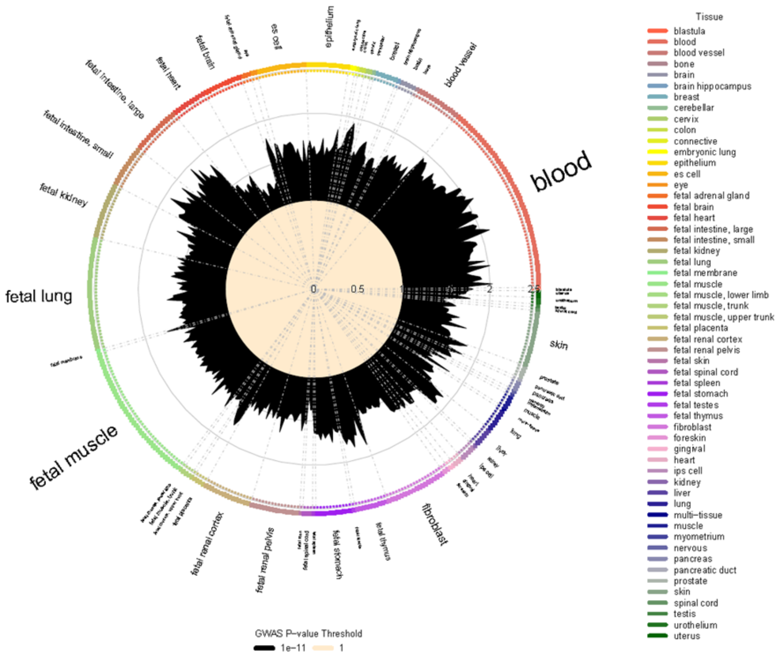

Peaks

H

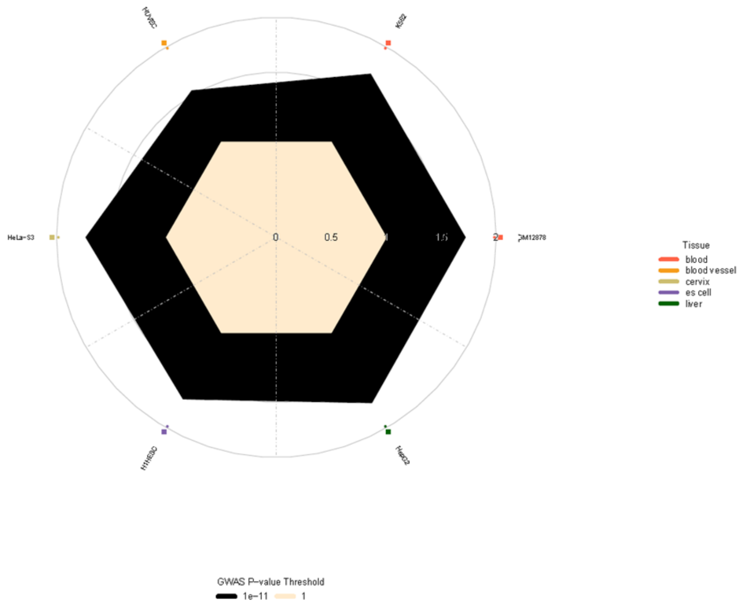

TFBS

B) African ancestry

A

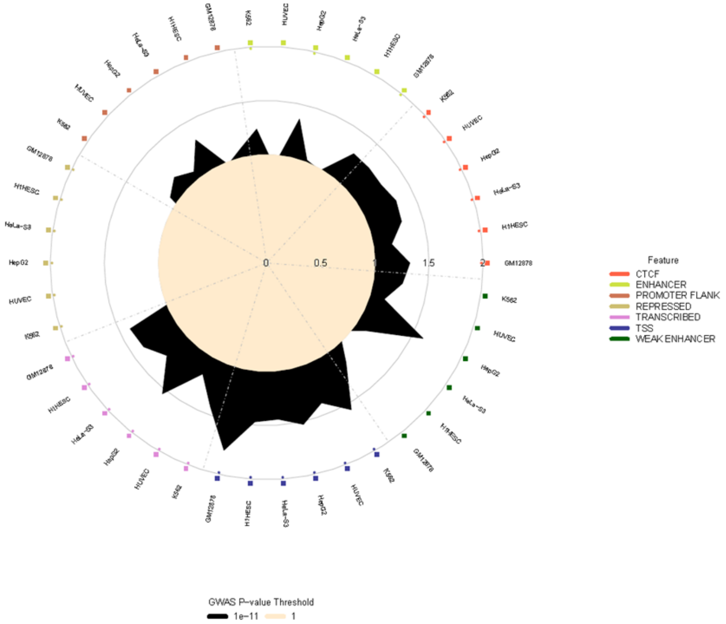

B

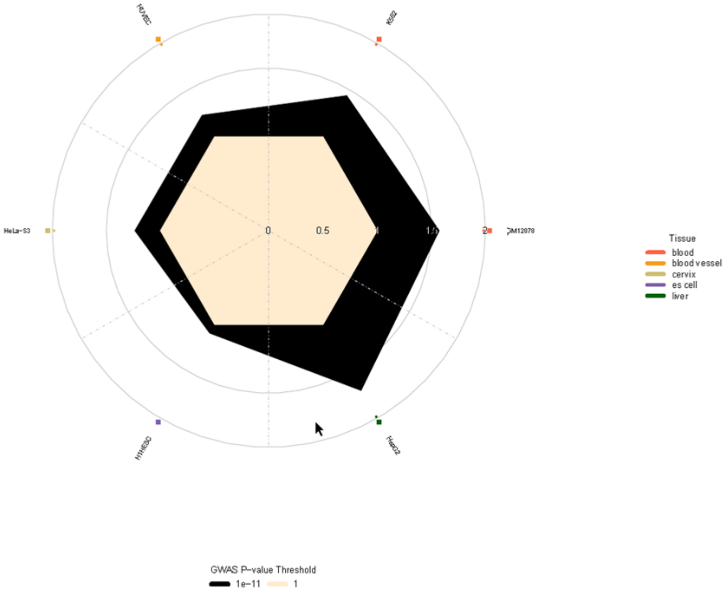

Chromatin States

FAIRE

C

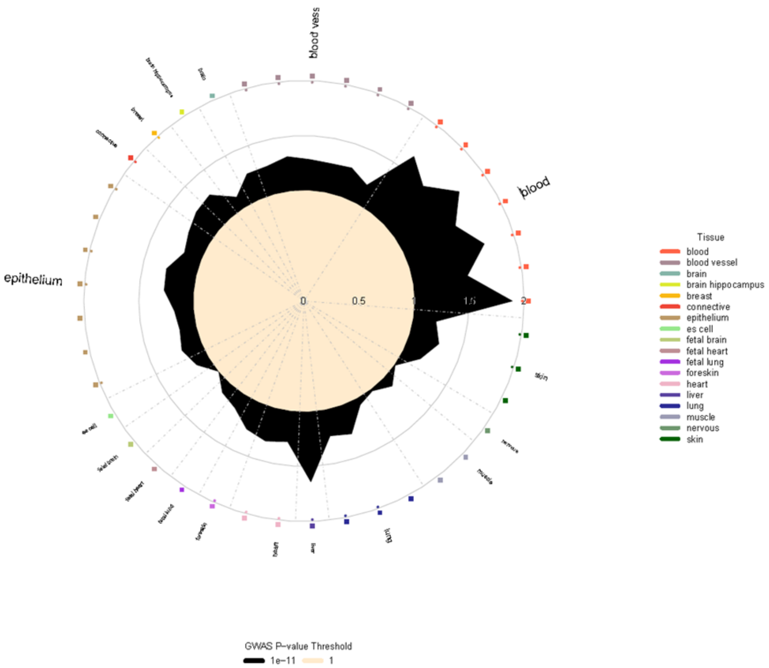

Footprints

D

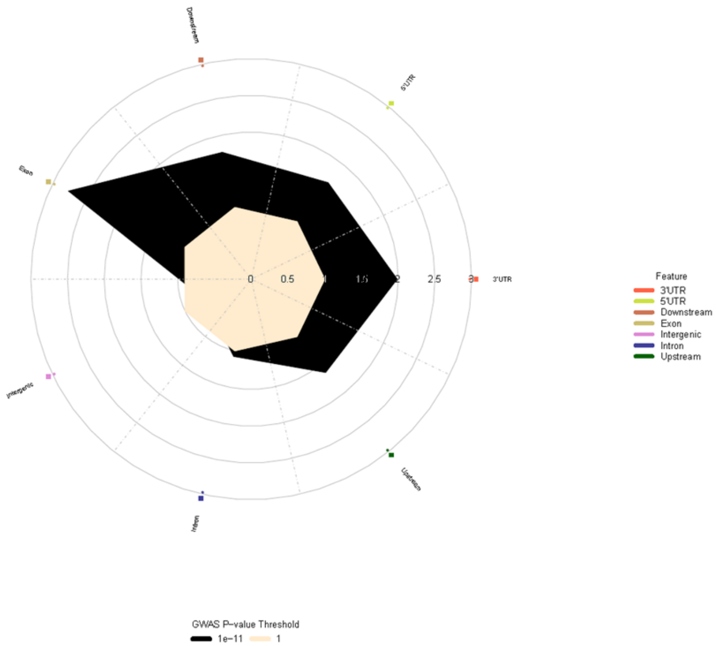

Genic

E

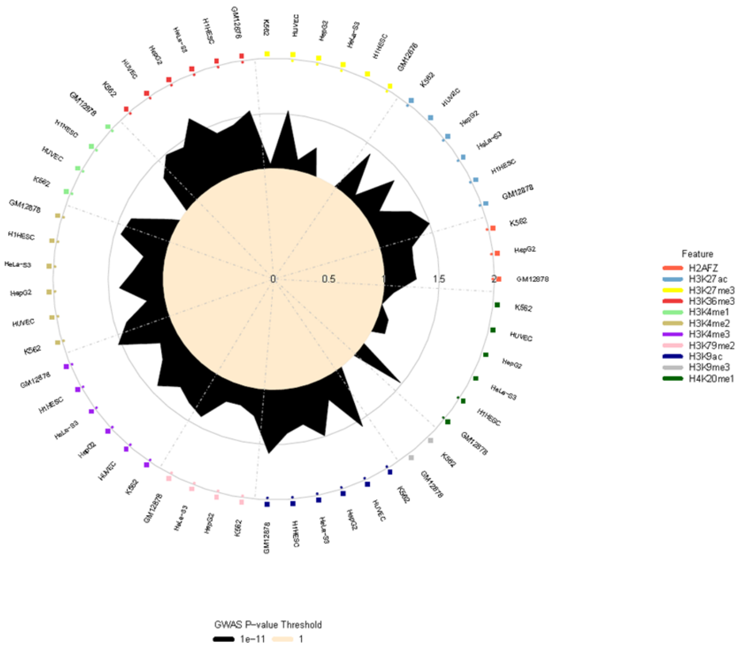

Histone Modifications

F

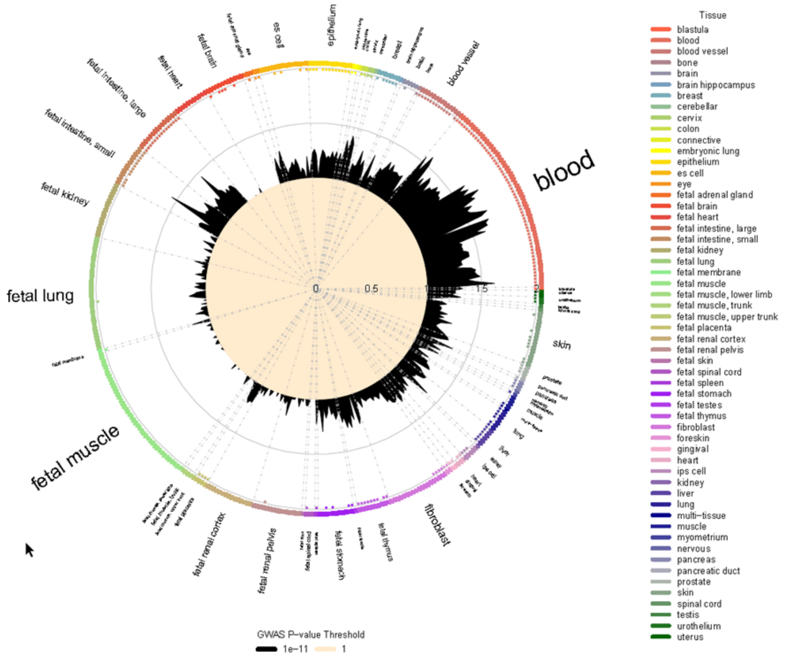

Hotspots

G

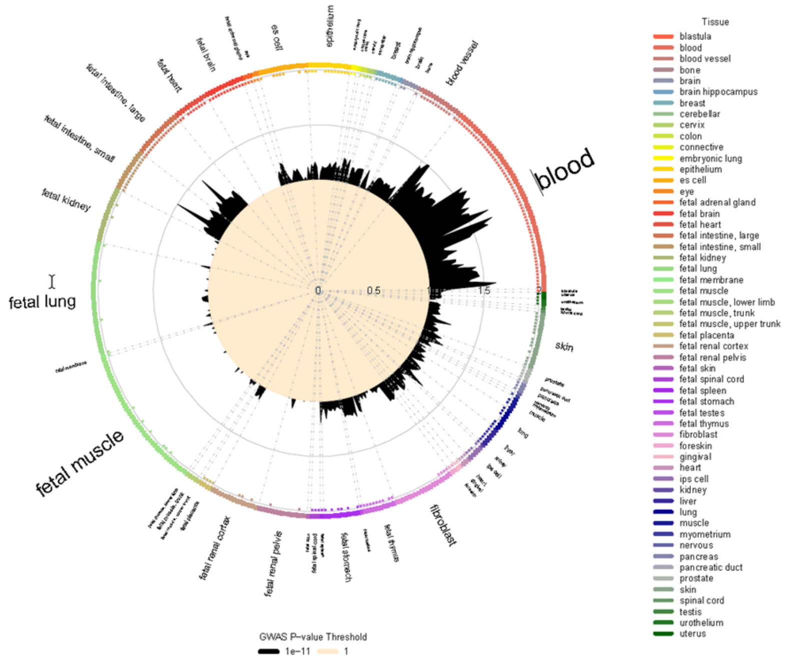

H

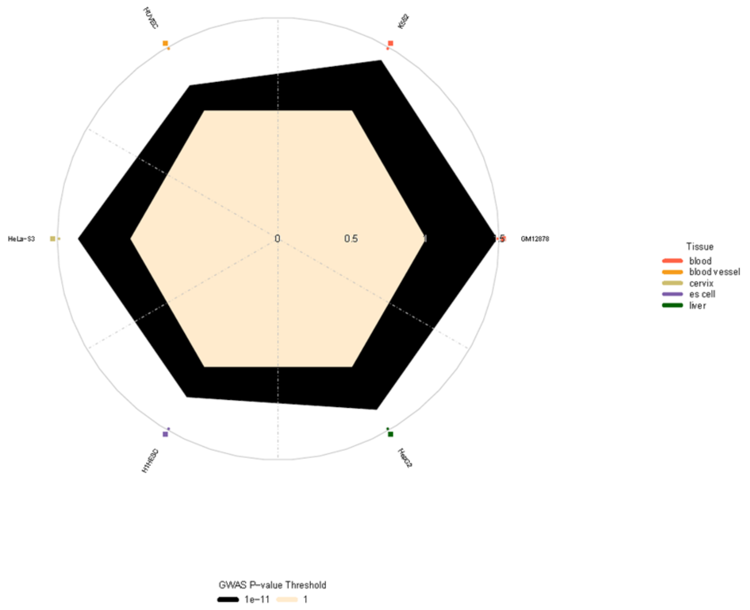

Peaks

TFBS

**Supplementary Figure 2.** Chord plots showing association of a genetic variant with proteins by ancestry groups for two examples of pQTLs classified as horizontal or vertical pleiotropic

A) An example of horizontal pleiotropy for rs3733402 on chromosome 4 (4q35.2)

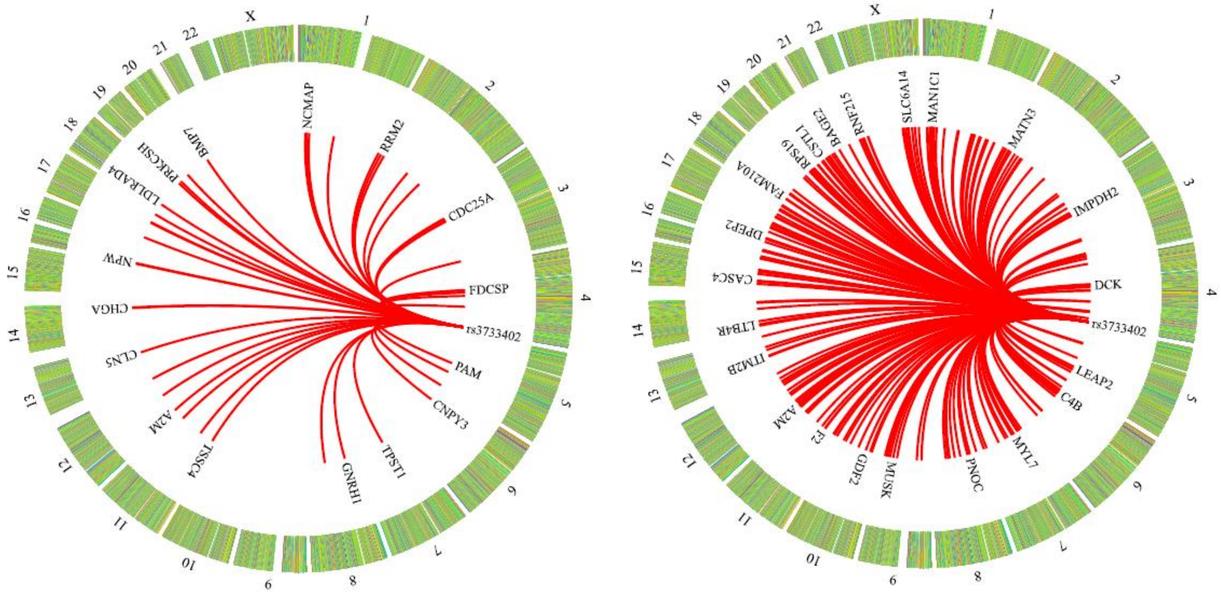

Lines link the genomic locations of rs3733402 and genes encoding their respective significantly associated proteins for AA (left) and EA (right). For clarity, only a small number of proteins were labeled.

B) An example of vertical pleiotropy for rs5471 on chromosome 16 (16q22.2) in AA

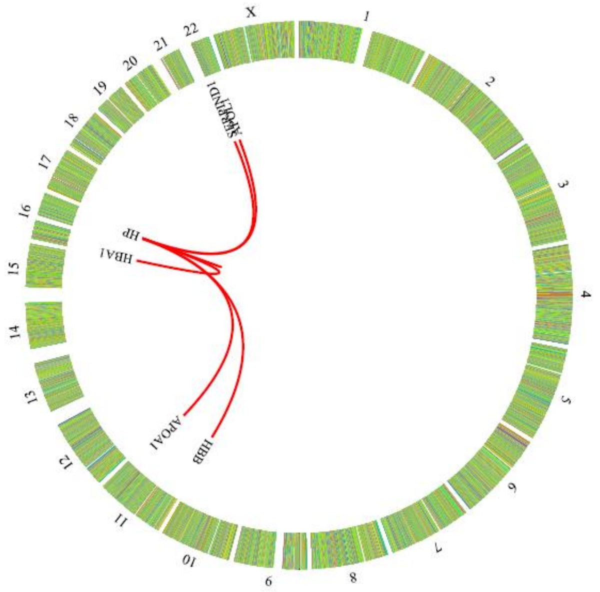

#### **Supplementary Figure 3**. Colocalization of pQTLs and genome-wide association signals of phenotypes.

Posterior probability of colocalizations of Heart Failure (HF) with pQTLs identified from EA and AA. Candidate causal variants for shared colocalization signals were marked with the SNPs' names.

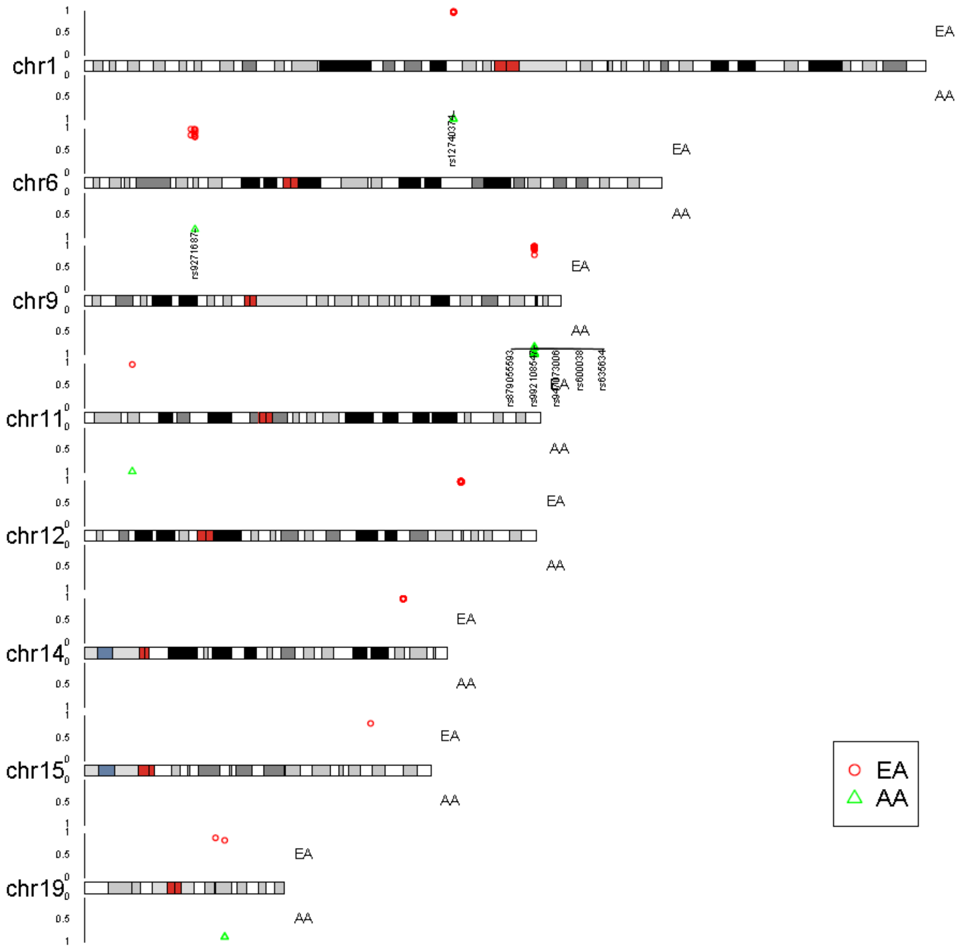

**Supplementary Figure 4**. Miami plot for results of Mendelian Randomization (MR) analysis of potential effects of plasma proteins on human phenotypes.

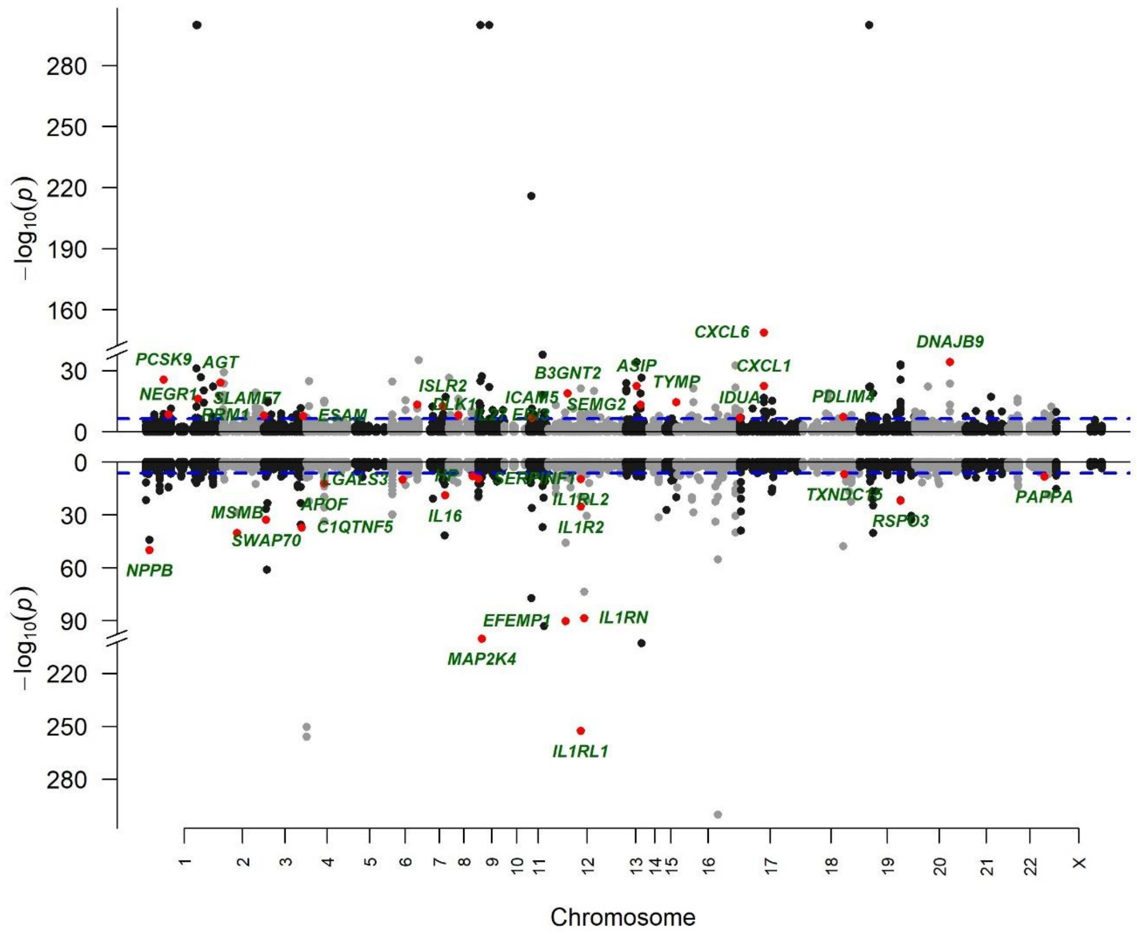

Each of the tested protein-phenotype pairs is represented by a filled circle. On the x-axis is indicated the chromosomal position of the coding gene of each protein and on the y-axis is indicated the -log10 p-value of the Mendelian Randomization (MR) results. Positive and negative effects of proteins are shown on top and bottom, respectively. The blue dashed line represents the Bonferroni significant level at $P<3.62\times10^{-7}$. Red color circles refer to the significant associations that were reported in phenome-wide proteomic MR mapping (1-3) and are labeled with gene symbols, where duplicated gene symbols were removed, keeping the one with a lower p-value.

#### Supplementary Tables

##### **Supplementary Table 1.** Descriptive table of 9,455 ARIC participants included in the proteomic GWAS analyses

| | **European ancestry** | **African ancestry** |
|---|---|---|
| **N** | 7,584 | 1,871 |
| **Male, N (%)** | 3,620 (47.7) | 715 (38.2) |
| **Age, mean (SD)** | 60.4 (5.7) | 59.2 (5.7) |
| **Age, median [25-75 pc]** | 60 [55-65] | 59 [54-64] |
| **Study Center** | | |
| **Forsyth County, NC (F)** | 2,273 | 226 |
| **Minneapolis Townships, MN (M)** | 2,798 | -- |
| **Washington County, MD (W)** | 2,513 | -- |
| **Jackson City, MS (J)** | -- | 1,645 |

**Supplementary Table 2.** Numerical break out of the associated regions after conditional analysis by ancestry groups

| | **African ancestry** | | | | **European ancestry** | | | |
|---|---|---|---|---|---|---|---|---|
| | **Total Number** | **Cis only** | **Trans only** | **Both** | **Total Number** | **Cis only** | **Trans only** | **Both** |
| **Sentinel variant to protein associations** | 1746 | 643 | 1103 | N/A | 4315 | 1050 | 3265 | N/A |
| **Genomic regions with pQTLs** | 807 | 567 | 199 | 41 | 1490 | 896 | 492 | 102 |
| **Proteins with pQTLs** | 1408 | 542 | 778 | 88 | 2565 | 571 | 1545 | 449 |

**Supplementary Table 9.** Numerical break out of the shared/validation status of AA aptamer-region pairs pQTLs in EA

| Type* | cis/trans | pQTL | Aptamer | UniProt | EntrezGene Symbol | Sentinel_variant |
|---|---|---|---|---|---|---|
| Shared | trans | 685 | 645 | 635 | 634 | 218 |
| Shared | cis | 428 | 428 | 418 | 417 | 421 |
| Validated | trans | 22 | 22 | 22 | 22 | 18 |
| Validated | cis | 14 | 14 | 13 | 13 | 13 |
| Not validated: NA | trans | 377 | 368 | 365 | 363 | 106 |
| Not validated: NA | cis | 189 | 189 | 184 | 184 | 184 |
| Not validated: Diff_nominal | trans | 9 | 7 | 7 | 7 | 8 |
| Not validated: Diff_nominal | cis | 4 | 4 | 4 | 4 | 4 |
| Not validated: Diff_Bonf | trans | 10 | 10 | 10 | 10 | 9 |
| Not validated: Diff_Bonf | cis | 8 | 8 | 8 | 8 | 8 |

* Shared indicates that the sentinel variant (or a proxy) validated at $P<1.025\times10^{-11}$. Validated indicates that the sentinel variant (or a proxy) was associated at the Bonferroni-corrected value ($P<2.86\times10^{-5}$). A pQTL identified in AA but not validated in EA was further categorized as the following three types:

(i) the sentinel variant or its LD-proxy was not included or did not pass the HWE P-value based filtering: NA

(ii) the validating variant (either the sentinel variant or its LD-proxy) had a P-value larger than the Bonferroni-corrected value: Diff_Bonf

(iii) the validating variant (either the sentinel variant or its LD-proxy) had a P-value larger than 0.05, or the effects in AA and EA were in opposite direction: Diff_nominal.

**Supplementary Table 10.** Numerical break out of the shared/validation status of EA aptamer-region pairs pQTLs in AA

| Type* | cis/trans | pQTL | Aptamer | UniProt | EntrezGene Symbol | Sentinel_variant |
|---|---|---|---|---|---|---|
| Shared | trans | 685 | 645 | 635 | 634 | 241 |
| Shared | cis | 428 | 428 | 418 | 417 | 419 |
| Validated | trans | 571 | 531 | 528 | 528 | 196 |
| Validated | cis | 150 | 150 | 148 | 148 | 150 |
| Not validated: NA | trans | 126 | 124 | 122 | 123 | 56 |
| Not validated: NA | cis | 87 | 87 | 84 | 84 | 85 |
| Not validated: Diff_nominal | trans | 680 | 620 | 614 | 610 | 379 |
| Not validated: Diff_nominal | cis | 158 | 158 | 157 | 157 | 157 |
| Not validated: Diff_Bonf | trans | 1203 | 983 | 970 | 969 | 447 |
| Not validated: Diff_Bonf | cis | 227 | 227 | 221 | 221 | 224 |

* Shared indicates that the sentinel variant (or a proxy) validated at $P<1.025\times10^{-11}$. Validated indicates that the sentinel variant (or a proxy) was associated at the Bonferroni-corrected value ($P<1.16\times10^{-5}$). A pQTL identified in EA but not validated in AA was further categorized as the following three types:

(i) the sentinel variant or its LD-proxy was not included or did not pass the HWE P-value based filtering: NA

(ii) the validating variant (either the sentinel variant or its LD-proxy) had a P-value larger than the Bonferroni-corrected value: Diff_Bonf

(iii) the validating variant (either the sentinel variant or its LD-proxy) had a P-value larger than 0.05, or the effects in AA and EA were in opposite direction: Diff_nominal.

**Supplementary Table 11.** Numerical break out of validation of pQTLs for conditionally independent aptamer-variant associations

| AA pQTLs | | | | | | |
|---|---|---|---|---|---|---|
| **cis/trans** | **Validated In EA** | **pQTL** | **Aptamer** | **UniProt** | **EntrezGene Symbol** | **Variant** |
| cis | Yes | 559 | 450 | 438 | 437 | 548 |
| cis | No | 412 | 333 | 324 | 324 | 397 |
| trans | Yes | 731 | 661 | 649 | 647 | 243 |
| trans | No | 458 | 425 | 422 | 419 | 157 |
| **EA pQTLs** | | | | | | |
| **cis/trans** | **Validated In AA** | **pQTL** | **Apatamer** | **UniProt** | **EntrezGene Symbol** | **Variant** |
| cis | Yes | 939 | 601 | 587 | 586 | 918 |
| cis | No | 1192 | 782 | 756 | 756 | 1151 |
| trans | Yes | 1367 | 1077 | 1058 | 1056 | 423 |
| trans | No | 2403 | 1613 | 1580 | 1574 | 1016 |

**Supplementary Table 12.** Numerical break out of replicated of previous pQTLs in AA and EA

| cis/trans | Replicated In EA | Replicated In AA | pQTL | Aptamer | UniProt | EntrezGene Symbol | Locus | Sentinel variant |
|---|---|---|---|---|---|---|---|---|
| cis | Yes | Yes | 432 | 420 | 410 | 410 | 424 | 427 |
| cis | Yes | No | 613 | 564 | 552 | 552 | 606 | 606 |
| cis | No | Yes | 77 | 76 | 76 | 76 | 77 | 77 |
| cis | No | No | 1468 | 1402 | 1372 | 1372 | 1444 | 1454 |
| trans | Yes | Yes | 905 | 808 | 798 | 798 | 282 | 290 |
| trans | Yes | No | 2986 | 1941 | 1902 | 1902 | 1115 | 1141 |
| trans | No | Yes | 79 | 73 | 73 | 73 | 47 | 48 |
| trans | No | No | 25346 | 4540 | 4352 | 4350 | 10448 | 11128 |

**Supplementary Table 17.** Overlap and colocalization of pQTL and eQTL pairs shared by EA and AA

| EntrezGene Symbol | EA_Sentinel | EA_Causal | AA_Sentinel | AA_Causal | LD_EA (r2) | LD_AA (r2) |
|---|---|---|---|---|---|---|
| *CLEC4C* | rs7302014 | rs7302014 | rs11055602 | rs11055602 | 1 | 0.891 |
| *PRSS57* | rs9304936 | rs9304936 | rs62131274 | rs62131274 | 0.909 | 0.11 |
| *GOLM1* | rs11141211 | rs11141211 | rs11141228 | rs11141227 | 1 | 1 |
| *GNLY* | rs12151742 | rs12151742 | rs12151742 | rs12151621 | 1 | 1 |
| *MDGA1* | rs10947693 | rs9349050 | rs9349050 | rs9349050 | 1 | 1 |
| *FOLR3* | rs71891516 | rs71891516 | rs71891516 | rs71891516 | 1 | 1 |
| *YWHAB* | rs6031847 | rs16989474 | rs4931 | rs4931 | 1 | 0.906 |
| *CPNE1* | rs56116518 | rs12480408* | rs147767406 | rs12480408* | 1 | 1 |
| *HIBCH* | rs291466 | rs291466* | rs291466 | rs291466* | 1 | 1 |
| *TP53I3* | rs10191964 | rs73920010* | rs1134516 | rs1134516 | 1 | 1 |
| *CBR1* | rs16993864 | rs16993864 | rs61034078 | rs16993864 | 1 | 1 |
| *TMEM106B* | rs3173615 | rs4721057* | rs4721059 | rs4721059* | 1 | 1 |
| *LIPN* | rs10509554 | rs10509554 | rs10509554 | rs10509554 | 1 | 1 |
| *QDPR* | rs67496406 | rs67496406 | rs67496406 | rs67496406 | 1 | 1 |

* In LD with missense variant in the gene encoding the target protein

**Supplementary Table 19.** Numerical break out of colocalized proteins and causal variants

| Ancestry group | Trait | N_protein | N_VarCisTran* | N_Variant** |
|---|---|---|---|---|
| EA | Sleep traits | 49 | 13 | 12 |
| EA | Others | 6 | 5 | 5 |
| EA | CVD traits - HF | 121 | 35 | 34 |
| EA | CVD traits - AF | 9 | 11 | 10 |
| AA | Sleep traits | 24 | 3 | 3 |
| AA | Others | 1 | 1 | 1 |
| AA | CVD traits - HF | 23 | 10 | 9 |
| AA | CVD traits - AF | 1 | 1 | 1 |

HF: Heart Failure, AF: Atrial Fibrillation

*N_VarCisTran: number of causal variants (a variant can be counted twice if it is causal to a protein as cis and is causal to another as trans)

**N_Variant: number of unique causal variants (a variant is counted just once no matter if cis or trans)

EA: European; AA: African

**Supplementary Table 22.** List of AA specific pQTLs not previously reported

| **SeqID** | **UniProt** | **Entrez Gene Symbol** | **Sentinel variant** | **RS_num** | **cis/ trans** | **Overlap With Sun** (4) | **Overlap with eQTL** | **Colocalized with eQTL** | **Included in Katz et al** (5) | **Included in Zhang et al** (6) |
|---|---|---|---|---|---|---|---|---|---|---|
| SeqId_8219_14 | O60844 | *ZG16* | chr16:51587827:T:C | rs2221099 | trans | Not Included | No | NA | NA | No |
| SeqId_9017_58 | P09848 | *LCT* | chr2:157269946:C:T | rs41416848 | trans | No overlap | No | NA | NA | No |
| SeqId_9017_58 | P09848 | *LCT* | chr2:122733333:C:T | rs1518056 | trans | No overlap | No | NA | NA | No |
| SeqId_9017_58 | P09848 | *LCT* | chr2:146265249:G:C | rs17736777 | trans | No overlap | No | NA | NA | No |
| SeqId_6577_64 | Q16363 | *LAMA4* | chr3:52800737:A:G | rs2710333 | trans | No overlap | No | NA | NA | No |
